# Tau accumulation patterns in PSP constrain mechanisms and quantify cell-to-cell and cell-autonomous aggregation rates

**DOI:** 10.1101/2024.12.14.24318991

**Authors:** Shih-Huan Huang, Annelies Quaegebeur, Tanrada Pansuwan, Timothy Rittman, Ruiyan Wang, Tuomas PJ Knowles, James B Rowe, David Klenerman, Georg Meisl

## Abstract

Protein aggregates are a hallmark of neurodegenerative disease, yet the molecular processes that control their appearance are still poorly understood. In particular, it is unknown to what degree the development of aggregates in one cell is triggered by nearby aggregated cells, as opposed to cell-autonomous processes. Here we develop a cell-level computational model to test alternative hypotheses of disease progression from human data and demonstrate its applicability in the primary tauopathy Progressive Supranuclear Palsy. From brain slices stained for aggregated tau, we quantify the contribution of cell-to-cell and cell-autonomous processes to the proliferation of aggregates across different brain regions and disease stages. We find that the triggering of aggregation by nearby aggregated cells, over distances in the order of 100µm, is the major driver of disease progression. Our computational model can then simulate interventions to evaluate potential therapeutic strategies in a virtual reconstruction of a human primary neurodegenerative tauopathy.

**Highlights:**

1. A minimal mathematical model can reproduce tissue-level aggregate accumulation patterns *in silico* at cellular resolution.
2. Cell-to-cell interactions determine aggregate patterns in progressive supranuclear palsy (PSP).
3. Cell-to-cell interactions are not limited to nearest neighbours, but act over a millimetre-scale.
4. Reducing cell-to-cell interactions or cell vulnerability, rather than targeting cell-autonomous processes, is a potential disease-modifying therapeutic strategy.

## INTRODUCTION

The association between misfolded, aggregating proteins and neurodegenerative diseases is well established^1,2^. Even though aggregating proteins are diverse and the aggregated structures they give rise to are specific to disease^3^, there are common principles of pathogenesis at the molecular and cellular level. These principles include the intrinsic ability of aggregating protein species to self-replicate^4^, their ability to overwhelm or avoid protein quality control and removal mechanisms^5^, and the potential to spread between cells^6–8^. It has been shown that both aggregate self-replication and spreading occur in animal models^9–11^. However, in most cases it has yet to be shown for human neurodegenerative diseases which is the critical – or rate-limiting – process. This is essential to guide therapeutic strategies to slow or arrest disease progression. A quantitative model of the molecular drivers of disease is required, that works at the spatiotemporal scale of human disease^12^.

Protein aggregation plays a central role across a spectrum of neurodegenerative diseases and has been studied in detail *in vitro* ^4,13,14^. Yet, the cellular and molecular processes found to control aggregate formation under such controlled conditions have proven difficult to relate to disease emergence and the pattern and pace of pathology observed in human disease^15^. Mathematical modelling of disease progression can bridge this gap, to link macroscopic patterns of progression to cellular and molecular processes. For example, models of selective vulnerability and connectivity can recover the brain-wide patterns of regions becoming affected in sequence over the course of different diseases^16^. Using such models, the rates of general classes of pathological processes have been quantified^10^, and the interaction of beta-amyloid and tau aggregation in Alzheimer’s disease (AD)^17^ has been elucidated. To date, these models of human disease have focused mainly on macroscale modelling, informed by whole brain imaging methods such as PET, MEG and MRI scanning^18–20^. This limits the conclusions that can be drawn about molecular and cellular mechanisms, and the applicability in drug development, which by its nature acts at the molecular level. New modelling approaches are therefore required that accommodate micro- and meso-scale processes.

With the advance of digital pathology and AI-facilitated automated classification of aggregated cell types^21^, cellular-level resolution of neuropathological changes can now be obtained and quantified at scale^21,22^. Such digital data with cellular resolution opens the door for models to investigate the rates of within-cell aggregation and cell-to-cell interactions.

Here, we present a model to link cellular aggregation patterns observed at the tissue level to underlying molecular processes. To achieve this, we use a minimal model of aggregation in a cell, and then allow cells to interact with each other to trigger aggregation. We apply this model to data from the primary tauopathy Progressive Supranuclear Palsy (PSP). Unlike Alzheimer’s Disease, PSP is associated with the aggregation of misfolded 4-repeat (4R) tau^23^, independent of the aggregation of a second misfolded protein (e.g. amyloid-beta in Alzheimer’s disease). PSP therefore provides an ideal test-bed for our approach, with its high clinicopathological correlation, and the high propensity of its 4R-tau to aggregate. Moreover, tau pathology in PSP follows a stereotypical sequential pattern, as described by Kovacs et al.^24^. We anticipate the lessons learned here will enable us to model Alzheimer’s Disease and other aggregation-related neurodegenerative diseases in the future.

We first introduce the mathematical model and demonstrate its behaviour when the different processes dominate. We then use an automated histological approach to capture the spatial aggregate patterns of histopathological images from 12 brain regions in 11 PSP cases, across disease stages^24^ (Table S1). The digitised information from nuclei and aggregated cell location is then used in our model to determine the relative contribution of molecular processes that gave rise to the observed patterns. This allows us to determine the rate-limiting steps in protein aggregation and progression, and investigate how the contribution of cell-to-cell and cell-autonomous processes to the proliferation of aggregates varies across brain regions and over the course of disease.

## RESULTS

### Modelling cell-level aggregate formation

A number of processes are needed to describe cellular level patterns of aggregate formation in neurodegenerative diseases such as PSP. The key processes of aggregate formation are: (i) *initiation*, de-novo formation of aggregates without the involvement of existing aggregates; (ii) *multiplication*, formation of new aggregates triggered by existing aggregates, for example via fragmentation; and (iii) *growth*, growth of existing aggregates by addition of further proteins. These processes couple together into a minimal reaction network that produces auto-catalytic amplification of aggregates, a feature observed for tau and across disease-associated proteins^4,25^. Protein synthesis and aggregate removal also play important roles in cellular aggregation, resulting in two distinct states for a cell: stable (healthy) and runaway aggregation (diseased)^5,26^.

In our model *in silico* system, we included the following core features: cells are in a stable state unless triggered to switch to a runaway aggregation state, either by a random cell-autonomous event, or by influence from other cells in the runaway aggregation state. This switch can occur when aggregate self-replication overwhelms clearance, or when significant amounts of preformed seed enter the cell^5^. Once triggered, a cell accumulates aggregates. There is evidence that aggregates are present at low concentrations in healthy states without triggering runaway aggregation^27^. This may represent a stable state in which aggregate production and removal are in balance^26^. We therefore assume that the large aggregate deposits visible in histological stains occur only in cells that have switched to the runaway aggregation state, and we refer to these as *aggregated cells* throughout.

Aggregated cells are capable of triggering aggregation in other cells. This could occur by transfer of aggregates via axonal connections that act as seeds, or by less direct means such as inducing inflammation^28^, Fig. 1A. The parameters of this model include a rate-constant for cell-autonomous triggering (*k_a_*), a rate-constant for cell-to-cell triggering (*k_s_*), and a characteristic length scale of cell-to-cell triggering (*σ*). As the cell-to-cell triggering process couples the behaviour of cells across space, we also refer to this process as spatial coupling.

**Figure 1:**
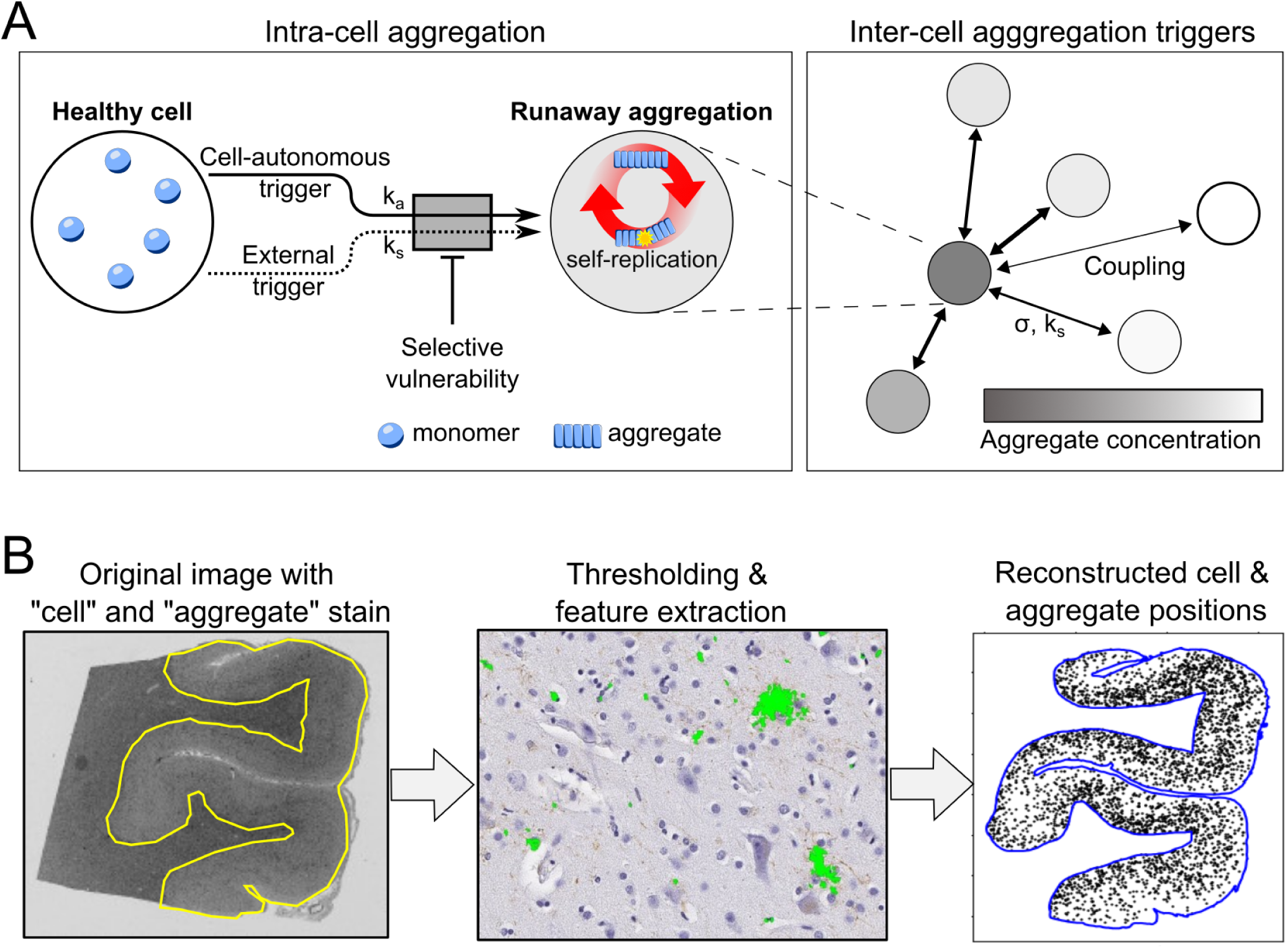
Schematic of the *in-vivo* cellular model for protein aggregation and our data porcessing pipeline. **(A)** In our model, the basic unit is a cell. Within each cell, once aggregation is triggered, aggregates rapidly accumulate. Aggregation can be triggered cell-autonomously (rate *k_a_*, units of inverse time), and additionally aggregated cells can trigger aggregation in other cells (as a model for e.g. transfer of seeds or indirectly mediated triggers), with a length-dependent coupling strength between cells (rate *k_s_* in units of inverse time, length dependence *σ* in units of length). How susceptible to being triggered a cell is, is determined by its *vulnerability*. As the simulation proceeds, we track the aggregation state of each cell over time. **(B)** Outline of the extraction of relevant information (nuclei and aggregated cell positions) from images of histopathological brain slices. Initially, only the grey matter is selected, then image segmentation detects aggregated cells and nuclei, and finally the cell position and cell state is reconstructed virtually. Scale bar in B: 5 mm (left), 50 µm (middle), and 5 mm (right).

We assume that the ability of an aggregated cell to trigger aggregation decays with distance as a normal distribution, with a standard deviation *σ* equivalent to the length scale of cell-to-cell triggering. A normal distribution describes the situation where triggering occurs by a randomly diffusing species, but may also be used as an approximation for how the average number of axonal connections varies with distance. As we show later, the exact choice of the functional form of the distance dependence does not affect our conclusions (Fig. S8). This description allows us to capture the different possible mechanisms for the evolution of the spatial aggregate patterns, often referred to as *spreading*. We avoid this term given the potential confusion as to whether it refers to the increase of the size of the region affected by pathology or the actual transfer of aggregated species.

Finally, to reflect recent biological insights^29,30^, we allow for a variable *vulnerability* term that defines the probability of a given cell being triggered, and reflects other biological process, such as different monomer expression levels or a varying ability to remove aggregates.

The behaviour of the model is demonstrated in Fig. 2 for a simple cell arrangement and under a variety of conditions. We will first discuss how to quantify the distribution patterns of aggregated cells that emerge from this model and how the different processes influence them. We go on to analyse data from PSP patients to quantify rates for the different processes modelled, and identify the rate-limiting steps. Finally, we use simulations on a virtual reconstruction of PSP brain tissue to illustrate how aggregated cell distributions evolve over the course of the disease, and to assess how varying the rates of disease processes would influence the outcome.

**Figure 2:**
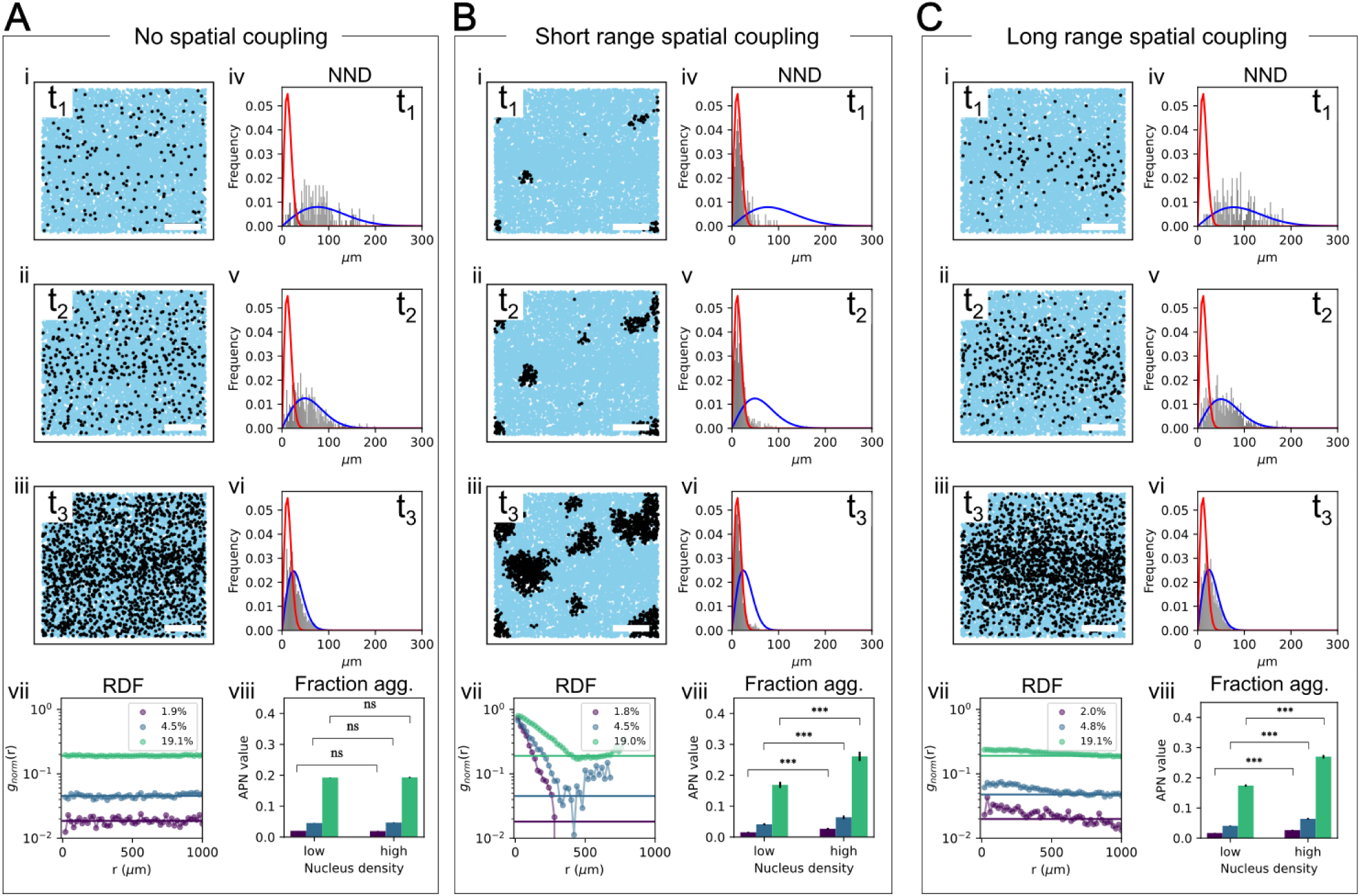
Simulation and analysis of intra-brain region patterns under varying spatial coupling conditions and fraction of cells aggregated. (A) Patterns observed with no spatial coupling across three fractions of aggregated cells: 2%, 5%, and 20%. (B) Patterns with short-range spatial coupling across the same fractions: 2%, 5%, and 20%. (C) Patterns with long-range spatial coupling across the same fractions: 2%, 5%, and 20%. In each panel, the top left sub-panels show 2D spatial patterns at different fraction aggregated, the top right sub-panels display the corresponding nearest neighbour distance distribution (grey histogram) with the analytical expression in the random limit (blue line) and the direct nearest neighbour coupling limit (red line), the bottom left sub-panels present the normalised radial distribution function (points) with the fully random distribution shown as a solid line (see Method for definition), and the bottom right sub-panels illustrate the aggregate-per-nucleus values in regions of varying nucleus density. Scale bar on panels i-iii: 500 µm. The simulation parameters for this figure are provided in *Simulation parameters*.

### Development of spatial measures to distinguish mechanisms

Brain slices from neurodegenerative disease contain a wealth of information, not just on the quantity of neuronal tangles and other deposits of aggregated protein, but also on the spatial distribution of these structures. This spatial distribution varies in different brain regions, and at different stages of disease. From the spatial distribution of aggregated cells it is possible to infer mechanistic information on the strength of cell-to-cell interactions, their length dependence and the importance of de-novo formation of aggregated structures compared to the formation triggered by nearby aggregated cells. However, extracting this information is non-trivial given the complexity of the images, the regional variation in brain structure (differences between brain regions, white and grey matter, cortical bands etc.) and the intrinsic stochasticity of the patterns. We therefore developed means to extract the relevant information from patient data.

As a first step, the histopathological images are processed, as detailed in the *Methods*, to automatically determine the position of aggregated cells and nuclei, Fig. 1B. These nuclei and aggregated cell positions are used in the remainder of this work; they can be directly compared to the output of our model. We now use our model to investigate different strategies to extract mechanistic information. A key property of the experimental data is that it captures only a single end-stage time point. This means that we will only be able to determine ratios of rates, rather than the rates directly. Nonetheless, this can still provide key mechanistic insight, as it is only the relative rate of processes that determines their importance in governing disease progression and thus makes them potential therapeutic targets. It is possible to estimate the absolute rate values from the duration of disease.

To decrease the complexity of the analysis process, we now discuss the results at two length scales separately. We will first demonstrate the measures developed on simulated data and then move on to applying them to patient data.

#### Nearest-neighbour distributions to investigate interactions between adjacent cells

We first focus on the fraction of aggregated cells in the immediate neighbourhood of an aggregated cell, which can be assessed at the shortest length scale reliably accessible from histopathological images. We use a local measure in the form of the nearest neighbour distance (NND), which is the distance from one aggregated cell to the next nearest aggregated cell, and compare this to the NND for all cells. Determining the NND for all aggregated cells or nuclei in a brain slice yields a NND distribution.

In Fig. 2 (panels i-iii in each condition) we have simulated a number of different scenarios for a simple arrangement of cells (random distribution of cells, with a denser band running through the centre). In Fig. 2 (panels iv-vi for each condition) we show the resulting NND distributions, which differ between scenarios permitting mechanistic conclusions to be drawn form the NND distribution. The most pronounced differences between scenarios are expected when few cells contain aggregates; as the fraction of aggregate cells increase the NND distributions for different mechanisms naturally converge as they become fully aggregated. We derived analytical expressions for the distribution of aggregates in the limits no spatial coupling and short range spatial coupling, which are shown superimposed on the example distributions in Fig. 2.

Briefly, when the appearance of new aggregated cells is spatially random with no triggering by other aggregated cells, the NND distribution of aggregated cells is much higher than the NND of all cells (Fig. 2A.iv-vi). At the other extreme, when the new aggregated cells are formed predominantly by an aggregated cell triggering its direct neighbours the NND distributions of aggregated cells closely resemble the NND distributions of all cells (Fig. 2B.iv-vi). By contrast, when cell-triggering acts over longer distances, the NND of aggregated cells again resemble the case where no triggering takes place (Fig. 2C.iv-vi). The NND distributions are thus a simple measure for the amount of cell-to-cell triggering occurring over short length scales, between directly adjacent cells. More advanced measures, such as the radial distribution function discussed below, provide additional information at longer length scales.

#### Millimetre length scale features can distinguish mechanisms

The analysis of NND distributions described above cannot distinguish the case in which cell-to-cell coupling is important but long-range (Fig. 2C) from one in which formation of aggregated cells is purely random via a cell-autonomous process (Fig. 2A). While the short-range distributions are comparable in both cases (Fig. 2A,C panels iv-vi), at the mm length scale the difference in spatial feature size is clearly apparent (Fig. 2A,C panels i-iii).

To quantify the spatial features at these longer length scales, we select two measures: the radial distribution function (RDF) and the locally averaged Aggregated cells Per Nucleus (APN) value. The APN is defined as the local fraction of aggregated cells as a function of the local cell density. The radial distribution function is a commonly used measure to quantify spatial distributions. By contrast, the APN value was developed in this work to help interpret the features of the aggregated cell distribution, exploiting variation in the underlying distribution of nuclei. A plot of APN vs cell density encodes mechanistic information, on how different cell densities affect aggregate formation. Cell densities vary over a mm length scale in brains, for example due to the presence of cortical layers, which can be exploited to gain mechanistic insights.

The APN value represents a fraction of aggregated cells, rather than an absolute number. This means it does not capture spatial features due to variations in cell density in a system where the rate of aggregate formation is unaffected by cell density. This is the case for example when only cell-autonomous formation takes place. However, when cell density affects aggregation, for example when cell-to-cell triggering depends on the separation of cells, we expect that these features will be captured in an APN vs nucleus density plot. Indeed, this is confirmed by our simulations: when cell coupling is important, denser regions show a higher fraction of aggregated cells due to the stronger coupling between the more closely packed cells (Fig. 2B,C, panels viii); whereas if there is no cell coupling, the APN values are the same in regions of different density (Fig. 2A.viii).

The RDF measures relative density along the radial axis. In its usual physical application, the RDF is normalised by the overall density such that an RDF value of 1 at a particular distance reflects a random arrangement. In our case we instead normalise the RDF to the overall density of cells, rather than the overall density of aggregated cells. This means the value of the RDF now denotes the fraction of cells aggregated at a particular distance from another aggregated cell. We also include the RDF expected in a random arrangement, the horizontal line, to highlight the degree of clustering compared to a random arrangement. In the no spatial coupling case we find, as expected, that the measured RDF is constant and overlaps with the RDF for a random arrangement (Fig. 2A.vii).

By contrast, in the short range coupling case, we find that the RDF peaks at short distances, with values close to 1 denoting fully aggregated regions (Fig. 2B.vii). The cluster size of densely aggregated cells is also visible from the extent of the peaks to hundreds of µm in Fig. 2B.vii. In the long-range spatial coupling case, the RDF still clearly peaks above the RDF of a random arrangement. The effect is much less pronounced than in the short range coupling case; even the centre of clusters are not close to being fully aggregated. The different conditions show clearly distinct patterns in the RDF, allowing us to infer mechanisms.

#### Cell types and vulnerability

In the simplest model all cells are assigned the same vulnerability, i.e. the same resistance against being triggered to aggregate. This is the assumption used throughout the majority of this work. However, to account for the fact that a number of different cell types are involved in the accumulation of aggregates and that there is increasing evidence for further heterogeneity in the vulnerability within a given cell type^31^, we also explored the effect of varying the vulnerability of individual cells (Fig. S9). We tested two types of vulnerability distributions: 1) at the one extreme we use a Bernoulli distribution where a cell can only have one of two vulnerability values, high or low; this could account for the presence of different cell types. 2) at the other extreme we use a uniform distribution, to model a continuum of vulnerability in the system. Our assumption of vulnerability imposed on individual cells does not depend on cellular location.

The results from simulating different vulnerability distributions show no qualitative changes in the NND distribution, RDF, or APN values at given fractions of aggregated cells. This means that conclusions about the dominant mechanism remain robust even when vulnerability distributions are not modeled in detail. The length scale of the cell-to-cell coupling, σ, appears to be weakly coupled to the vulnerability, thus the absolute values of this parameter are expected to be less accurate in the absence of an accurate measure of cell vulnerability variation.

Having established the spatial features in simulation, we next consider post-mortem data from people with PSP.

### Mechanistic information from PSP brains on different length scales

The aggregates are identified from the brain slice images by image analysis and then classified into different subtypes, tufted astrocytes (TA), which are formed in astrocytes, coiled bodies (CB), which are formed in glial cells, and neurofibrillary tanlges (NFT), which are formed in neurons, by a machine learning algorithm^22^. Their positions, as well as all nucleus positions, are recorded and will be used in the subsequent analysis. This process is performed on all images, across stages and brain regions. For details see Methods.

#### Random distribution dominates at cell-level length scales

The NND distributions show that there is little coupling between directly adjacent cells and the RDFs show little variation on the length scale of ∼ 100 µm, regardless of brain region and stage of the disease (Fig. 3D, I, N, and 4A). This finding rules out a mechanism where an aggregated cell simply triggers aggregation only in its closest neighbours (Fig. 2B). Therefore mechanisms that are expected to transfer aggregates only to directly neighbouring cells to induce aggregation there, such as tunnelling nanotubes^32^, are unlikely to be dominant processes. This leaves two other possibilities: The simplest is that appearance of new aggregated cells is fully cell-autonomous, also at longer length scales, and cells become aggregated independently of any aggregated cells in their vicinity (Fig. 2A). This explanation of course would be somewhat at odds with the hypothesis of seeding, i.e. the ability of preformed aggregated tau to induce the aggregation in new cells, observed in many model systems, as well as the observation of exponential amplification of tau concentrations in disease. The other mechanistic interpretation is that inter-cell transmission does happen, but the ability of an aggregated cell to induce aggregation in other cells is not limited to those cells close by and instead decays only slowly with distance from the aggregated cell (as in the simulated example in Fig. 2C). In this scenario aggregated cells are still responsible for triggering the aggregation, but at short length scales this *aggregation pressure* is essentially spatially uniform and the location of newly aggregated cells is governed by stochastic effects, such as the cells’ differing vulnerabilities to become aggregated.

**Figure 3:**
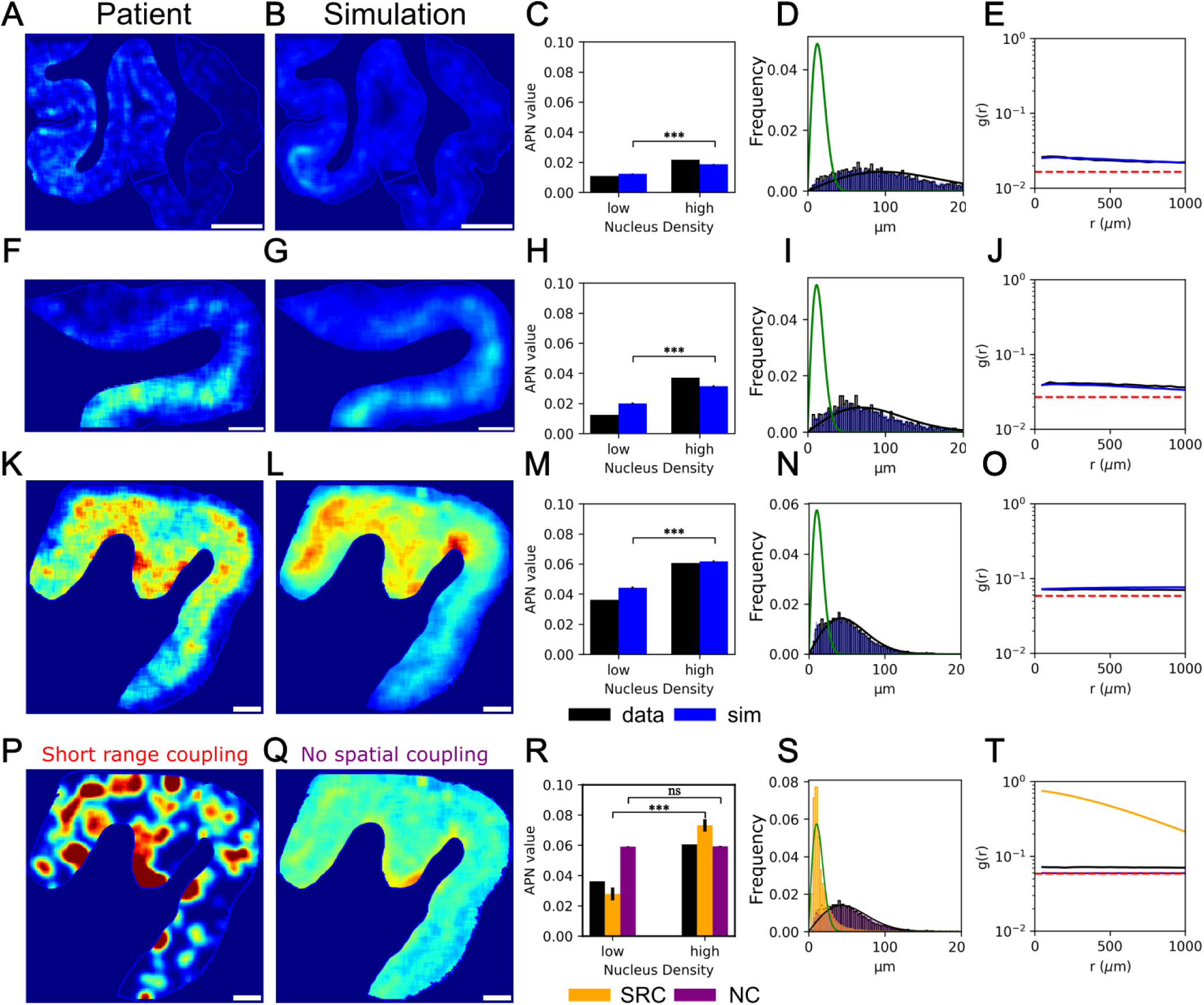
Comparison between real brain data and simulation results. Panels (A)–(O) show comparisons between real brain data and simulations across different brain regions and the disease stages defined by Kovacs et al.^24^. (A), (F), and (K) display the rolling average aggregate density from real data, while (B), (G), and (L) show the corresponding simulations using the best fit parameters. (C), (H), and (M) show the aggregate-per-nucleus (APN) values in both low and high nucleus density regions, for the patient data (black) and the simulations (blue, mean of 10 simulations, error bar is standard error of the mean). (D), (I), and (N) show the nearest neighbour distance distributions for the patient data (black histogram), the simulations (blue histogram, from a single simulation), the analytical expression in the random limit (black line, see Method for the derivation) and the direct nearest neighbour coupling limit (green line, see Method for the derivation). (E), (J), and (O) show the radial distribution function for the patient data (black) and simulations (blue, from a single simulation) with the fully random RDF shown in dashed red. The patient data are from the temporal cortex at Kovacs stage 3 (A-E), premotor cortex at stage 4 (F-J), and primary motor cortex at stage 6 (K-O). Panels (P) and (Q) show two simulation misfits to the patient data in (K), with the coupling radius set to 1*/*8 of the best fit value and the ratio *k_s_/k_a_* set to 10 times the best fit value in (P), coloured red in (R-S) and the cell-to-cell coupling switched off while leaving the other parameters unchanged in (Q), coloured purple in (R-S). (R) compares APN values, (S) compares nearest neighbour distributions, and (T) compares radial distribution functions between the misfits and data. Scale bar = 2 mm.

**Figure 4:**
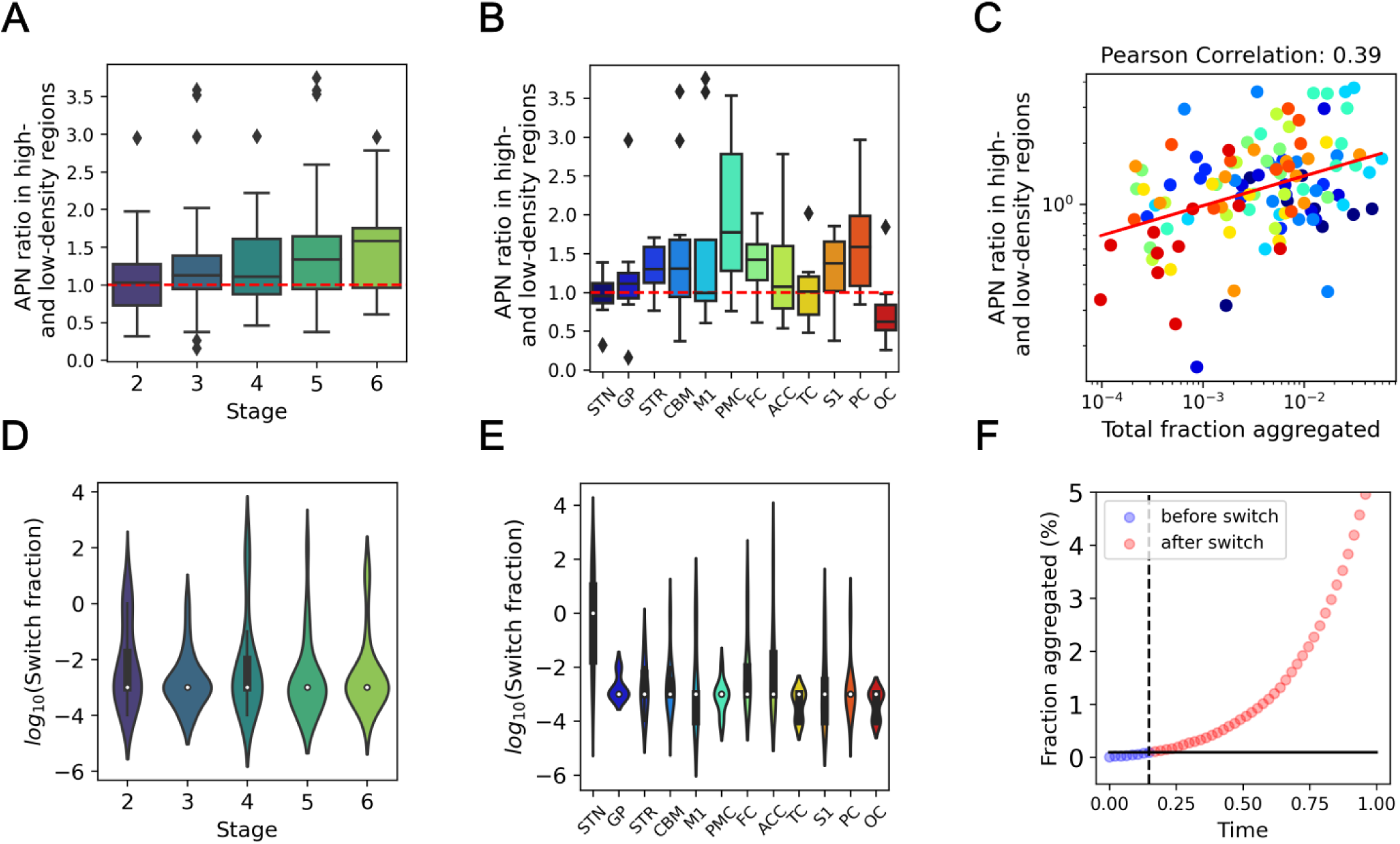
Model-free analysis and fitted parameters from all patient data. (A) Box plots summarizing the ratios in APN values across different nucleus density regions for each stage. (B) Box plots summarizing the ratios in aggregate-per-nucleus (APN) values across different nucleus density regions for each brain region. For (A) and (B), the box represents the interquartile range (IQR), encompassing the middle 50% of the data with edges at the first and third quartiles. Whiskers extend to 1.5 times the IQR from the quartiles to show the data range, while points outside these whiskers are plotted as outliers. (C) Correlation between the APN ratio in high- and low-density regions against the fraction of aggregated cells. The red line represents the fitted linear curve for the scattered points. Each point is color-coded by brain region, using the same color scheme as shown in panel (B). (D) Violin plots summarizing the switch fraction for each stage. (E) Violin plots summarizing the switch fraction across different brain regions (F) Fraction of aggregated cell over time, from onset (no aggregates) up to the fraction aggregated seen in stage 6 (≈ 5% aggregated). Circles denote simulations based on the cell positions from the brain slice shown in Fig. 3K. The solid black line is switch fraction, the dotted black line is the time when the simulated fraction aggregated reaches switch fraction. Blue and red denote times before and after switch time, respectively.

To better understand the pathogenesis at short distances we also investigated the NND of aggregated cells in different aggregated cell types separately (TAs, CBs and NFTS). The distributions resemble those observed for all aggregated cells, consistent with the previous mechanistic conclusions (Fig. S5A-C). Finally, we computed the nearest neighbour distance across different aggregated cell subtypes, that is the distance between for example tufted astrocytes and the nearest coiled body (Fig. S5D-I). We can compute the average NND for a random arrangement of cell types and a random appearance of aggregated cells. If there was increased triggering from one type to another, for example if most NFTs were formed by triggering from a TA, then we would expect a lower than predicted cross-NND in the corresponding plot. In practice, we find that the average NND between two different types is slightly larger than the prediction for a random arrangement of cell types and aggregated cells (i.e. above the line of equivalence in Fig. S5). This indicates that specific arrangement of cell types, not appearance of aggregated cells, dominates these patterns and there is no evidence for preferential coupling from one cell type to another.

#### Intra-brain-level aggregated cell distributions are not consistent with cell-autonomous triggering but imply long-range coupling

While nearest neighbour distributions are a good measure for the spatial distributions at short length scales, to quantify the aggregated cell distributions at the longer mm-level length scales, more complex measures that can quantify spatial features need to be employed.

In Fig. 2 above we showed that the locally averaged fraction of aggregated cells (or APN value) and radial distribution function (RDF) are two measures that display different features depending on the mechanism that dominates the appearance of aggregated cells. In Fig. 3A, F and K we show the APN value across a brain slice, for 3 example datasets at different disease stages. In Fig. 3C, H and M we show the corresponding average APN value in high and low density cell regions. A more detailed plot, for these and additional datasets, is shown in Fig. S6. In all of these images, the APN value is clearly higher in the dense cell regions than the less dense cell regions. This observation cannot be explained with a simple cell-autonomous model, even when allowing for cell density-dependent vulnerability as discussed below. The observation is however consistent with a simple cell-to-cell coupling model, where the coupling two cells is stronger the closer together they are.

The behaviour of the RDF is also in agreement with these conclusions based on the APN value. The RDF values are significantly above the value for a random arrangement (dashed red line in Fig. 3E, J, and O). This suggests that cell-to-cell coupling plays a significant role at longer length scales, reinforcing the idea that aggregation is not merely a local event but is influenced by spatial interactions extending beyond immediate neighbours. We now move beyond these qualitative observations to extract the parameters of our model that best describe the data by fitting.

#### Estimating parameters from the data

Using our model we can identify which molecular mechanism dominates in human disease, and quantify the rate constants that best match the patient data. In order to do so we use the cell positions given by the histopathological images to reconstruct a virtual brain slice *in silico*. We can then run simulations on this specific cellular distribution to determine which set of parameter values best describe the experimental data. This allows us to determine the relative importance of cell-autonomous over cell-to-cell triggering processes, given by the ratio of the cell-autonomous triggering rate and the rate-constant for cell-to-cell triggering *k_a_/k_s_*, as well as the spatial coupling radius *σ*.

APN histograms, rather than the full 2D images, were used to evaluate the match of simulations to data. This is motivated in part by the intrinsic stochasticity to the aggregation process: which cell will be triggered, either by cell-autonomous processes or cell-to-cell coupling, is to some degree a random process. This means that individual realisations of each simulation will yield slightly different results (Fig. S7). This reflects the biological situation and similar stochasticity would be observed in the experimental data if it were possible to produce an exact repeat. In practice this means that simulations are unlikely to match the data cell-for-cell even if the model fully captured all biological processes taking place. However, higher level features, such as those described by the NND distribution, the RDF or the APN value for different cell density regions, are less affected by this stochasticity and therefore better suited for inference. We use the APN histogram here, but other measures, such as the radial distribution function, can also be used. The results of using RDF for inference are consistent with those presented here and are shown in the supplementary information (Fig. S12).

As shown for the example images in Fig. 3, the best fit simulations match the data not only at the level of the spatial measures of APN value in high and low density regions, NND distributions and RDF, but also surprisingly well at the 2D image level, see Fig. 3 B,G and L compared to Fig. 3 A, F and K. Some discrepancy between the empirical data and the best fit simulations at the level of the 2D images in Fig. 3 is due to stochasticity, as discussed above. We also recognize that our model may not capture all the intricacies of the spatial dynamics of aggregation and cell interaction. However, it can explain the major features of the data well, with the best fit rate constants being consistent across samples (further details see next section). To put into perspective the goodness of fit, we show two misfits in Fig. 3 P-T. In Fig. 3P we have forced the coupling radius to be 100 *µm* and the rate ratio of cell-to-cell trigger and cell-autonomous trigger to be 10000 (compared to a best fit value of 800 µm and 1000), whereas in Fig. 3Q aggregation is triggered only by the cell-autonomous process and there is no cell-to-cell coupling. The discrepancy between the misfits and the empirical data is clear across all measures, highlighting that neither can explain the experimentally observed data.

#### Trends across disease stages and brain regions

In Fig. 4 we summarise the conclusions across stages and brain regions for our dataset, which includes 11 patients, each with up to 11 brain regions (detailed numbers see Tab. S1). We show both the results of a direct, model-free analysis of the data, Fig. 4A-C, and of the model best fits, Fig. 4D, E.

We find that the average NND is close to that predicted when there is no coupling between nearest neighbours, at all disease stages and in all brain regions, see Fig. S13A. Zooming out to longer length scales reveals evidence for spatial effects. At mm length scales, there is significant variation in cell density, so the dependence of the APN value on cell density contains mechanistic information. Fig. 4A and B show the ratio of the APN value in high and low nucleus density regions, grouped either by stage (A) or by brain region (B). Values above 1 denote that cells in high nucleus density regions are more prone to aggregate than in low nucleus density regions. This is the case at all stages from stage 3 onwards and is most pronounced in stages 5 and 6. The same trend is observed for the RDF, see Fig. S13B. When grouped by brain region, most brain regions also display a higher aggregation propensity in denser cell regions, although errors are larger given the lower number of samples in each group. The STN is an exception, with APN ratio = 1, which might reflect biological distinctions of interest, or result from its very small volume and specific cellular conformation. The occipital lobe is the only region with APN ratio below 1. This may reflect its status as the last region to develop significant tau-pathology under the Kovacs staging system. Consistent with this hypothesis, we show in our more detailed analysis below that the small number of aggregated cells present in the OC is not sufficient for cell-to-cell coupling effects to be important.

In summary, the model-free analysis shows that aggregated cells in the immediate vicinity of a cell play little role in determining its aggregation state, yet on 100s of µm to mm the density and fraction aggregated of the surrounding cells has a significant influence. Given the apparent importance of cell density in determining aggregation propensity, we further investigated the potential mechanistic origins of this effect. There are two basic scenarios: either cells in dense regions more vulnerable to aggregate, or the vulnerability of dense regions is simply a result of the fact that it is easier for cells to couple when they are close together. The ratio of the APN value of low and high cell density regions at different disease stages can answer this question: if the effect of high cell density regions were to simply increase the vulnerability of cells, and there was no cell-to-cell coupling effect, one would expect high cell density regions to be more aggregated at any fraction aggregated. By contrast, if the effect was due to the easier triggering of aggregation between closely packed cells, rather than an increased vulnerability in the dense regions, we would expect the difference in APN value between high and low density cell regions to become more pronounced the more aggregated the system is. This is because at low fraction aggregated, cell-to-cell coupling is of lower importance so the differences between low and high cell density regions would not be as pronounced. By plotting the ratio of the APN value in the low and high cell density regions against the total fraction of aggregated cells, Fig. 4C, we see that the experimental data fall into the latter category. There is a noticeable increase in the difference between low and high cell density regions as more aggregated cells accumulate. In fact, under low aggregate conditions, there are relatively fewer aggregated cells in high cell density regions, implying that if there is a vulnerability difference between high and low cell density regions, high cell density regions are less, rather than more, vulnerable.

These findings support our choice of model for of a spatially uniform vulnerability and a cell-to-cell coupling determined by cell separation. Using this model to fit the data by matching APN histograms (See Method Sec. *Parameter inference* for fitting details), as outlined in the above section, we show that indeed the observed patterns can be matched. We thus obtain best fit values for both the relative importance of cell-autonomous over cell-to-cell triggering processes, given by the ratio *k_s_/k_a_* (Fig. S10), as well as the spatial coupling radius *σ* (Fig. S11, S12).

The ratio *k_a_/k_s_* is an estimate for the overall fraction aggregated at the point where cell-to-cell coupling begins to dominate over cell-autonomous trigger, which we refer to as the *switch fraction*. As derived in the methods, *k_a_/k_s_* is an approximate expression for the switch fraction. We show a simplified derivation of this quantity in the methods. Fig. 4D &E shows that the switch fraction across all stages and all brain regions is on the order of 0.001, or 0.1% of cells aggregated. Fig. 4C, shows that most brain regions have already exceeded the switch fraction at the time of measurement, with the exception of the OC, which is the last brain region involved in the disease progression. STN is a notable outlier in Fig. 4E, due to its early involvement in disease^24^. The resulting extensive cell death^33^ renders our models, which do not include cell death, unable to fully explain the patterns. In Fig. 4F we show the simulated accumulation of aggregated cells over time, from an aggregate free state, to one that has 5% of cells aggregated (corresponding approximately to the highest levels of aggregation observed in patient data), in a virtual reconstruction of a typical brain slice. Two phases can be identified, corresponding to the cell-autonomous phase before the switch fraction is reached, and the cell-to-cell phase after the switch fraction is reached. Note the much faster rate of increase in the latter parts of the cell-to-cell phase. We assume constant rates and vulnerability over time for this plot, in reality ageing effects may be important in particular in determining the time of onset and progression in the cell-autonomous phase, as discussed as part of the *limitations* section below.

These findings imply that it is very unlikely that a single cell-autonomous event triggers disease progression. Instead, they predict that many cell-autonomous aggregation events take place before cell-to-cell mechanisms become dominant when on the order of 0.1% of cells are already aggregated.

The change in dominant mechanism suggests that different therapeutic interventions may be effective at specific stages of the disease. When the aggregate fraction is below the switch fraction, therapies should target cell-autonomous mechanisms. Once the switch fraction is exceeded in a brain region, interventions should focus on cell-to-cell coupling mechanisms. Note however that some interventions, such as reduction of the tau concentration could affect both processes equally, and that in practice it may be hard to administer therapies early enough to affect the cell-autonomous phase. Below, we will demonstrate that targeting the wrong mechanism results in negligible effects on aggregate accumulation.

Our model fits also determine the coupling radius, i.e. the characteristic length scale over which cells can trigger aggregation on other cells. However, the current dataset does not provide strong constraints on its value. The coupling radius most consistent with the data is in the range of several hundred µm to 1 mm (Fig. S11 and S12), implying that cell-to-cell coupling extends well beyond nearest neighbours but still resulting in spatial heterogeneity within a brain region. Note, that this value was obtained under the assumption of constant vulnerability and a significant variation in vulnerability could lead to a lower value of the coupling radius. The fact that no independent quantitative measurement of vulnerability exists however means that we can only report an effective coupling radius for the assumption of constant vulnerability. By contrast, the switch fraction is independent of vulnerability, so its value is applicable also in the case of varying vulnerability.

### Simulating effectiveness of therapeutic strategies

Having determined the rate constants of the individual processes in the preceding section, we can now use these insights to simulate how aggregated cell distributions may have evolved in a virtual reconstruction of each brain. Crucially we can also investigate how the distributions would be affected if a therapeutic intervention to slow a specific process were administered at different stages of the disease. We illustrate this using the primary motor cortex of a stage 6 patient in Fig. 5. A common initial state, *t*_0_, corresponds to 1.8% fraction aggregated or Kovacs stage 3-4 (Fig S1A), from which we start simulations under 3 conditions: with the best fit rate constants determined from the patient data, Fig. 5A-D, with the cell-to-cell triggering rate lowered by 50%, Fig. 5E-H, with the cell-autonomous triggering rate lowered by 50%, Fig. 5I-L, and with the vulnerability lowered by 50%, Fig. 5M-P. The simulations are all run for the amount of time it takes the unaltered conditions to reach the late stage disease state observed in the patient data. A clear slowing in the accumulation of aggregated cells is achieved with a 50% reduction of either the cell-to-cell triggering rate or the vulnerability. By contrast, lowering the cell-autonomous triggering rate by 50% leaves the progression essentially unchanged. This highlights that the cell-autonomous triggering rate becomes essentially irrelevant for the overall rate of progression, and therefore a poor drug target, once a certain fraction of aggregated cells have formed. It also showcases how modelling and simulation can be used to investigate the effect of altering different microscopic processes.

**Figure 5:**
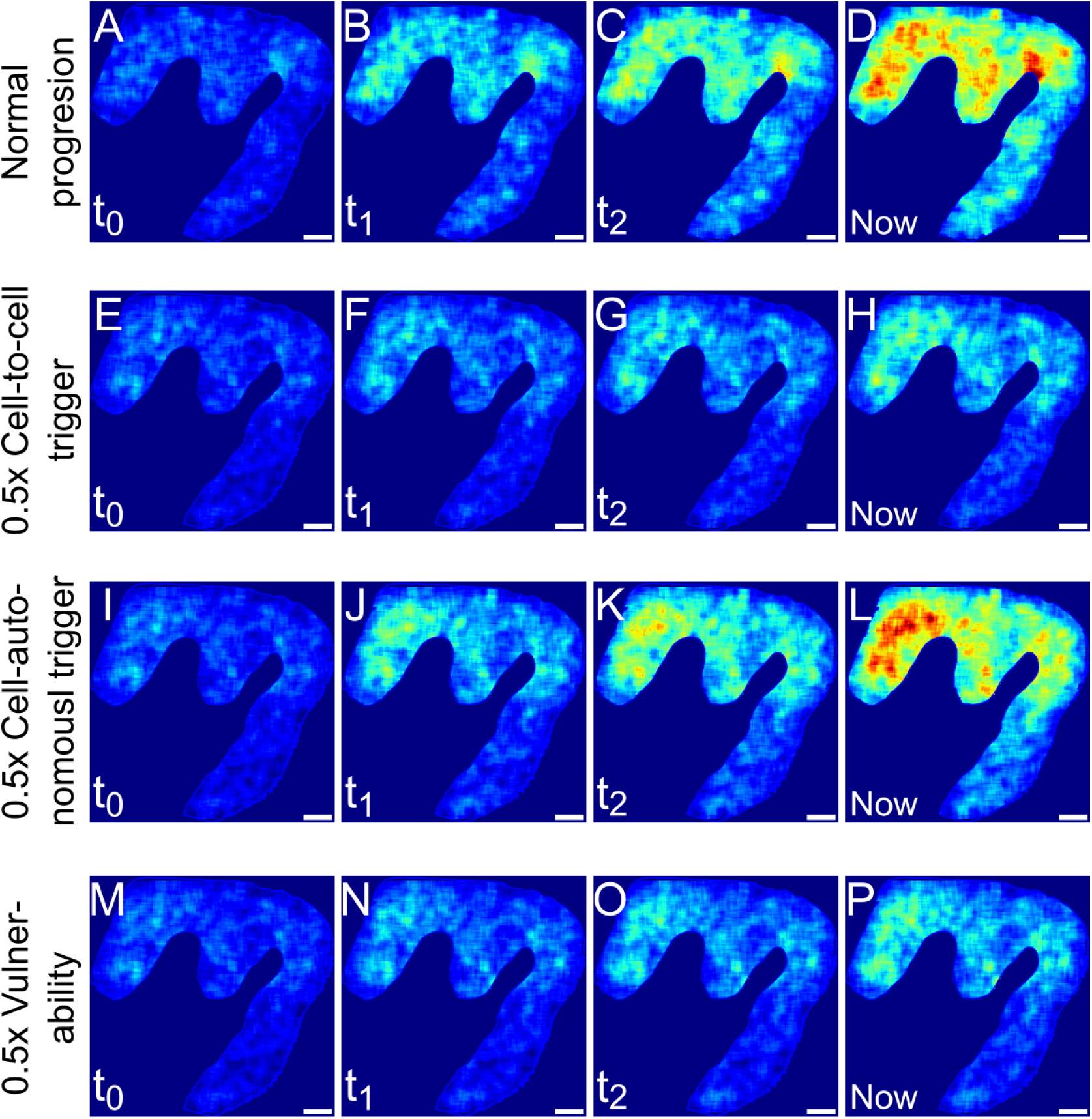
Simulations on virtual reconstruction of brain region. (A-D) A virtual reconstruction of cell positions from measurements of brain slices allows us to simulate how the aggregate distributions may have evolved since the onset of the disease. (E-H) Temporal dynamics of the simulation based on cell positions from (A) with a 0.5-fold rate of cell-to-cell triggering. (I-L) Temporal dynamics of the simulation based on cell positions from (A) with a 0.5-fold rate of cell-autonomous triggering. (M-P) Temporal dynamics of the simulation based on cell positions from (A) with a 0.5-fold of vulnerability. The same initial state at *t*_0_ is used in all conditions, *t*_1_ is half way between *t*_0_ and *Now*, *t*_2_ 71% of the way. Scale bar = 2 mm

### Limitations and future directions

There are several potential limitations and areas for optimization in our model. In particular, we focussed on developing a minimal model that captures the dominant features, thus the current framework does not explicitly include the effects of cell death, the detailed dynamics of intracellular aggregate formation and removal, the role of different types and sizes of aggregated species and the ageing processes. The current model can explain well the aggregated cell patterns seen, and infer the relative importance of general classes of mechanisms, such as cell-autonomous and cell-to-cell triggers. However, a detailed time-course and description of disease onset may require the effects of ageing to be included. Effects of ageing are likely to be reflected as an increase in the rates or in vulnerability in our model^34–37^. This would compress the time-course shown in Fig. 4F at later times and lead to a more sudden increase in the rate of aggregation, still consistent with late onset and rapid progression of disease. Furthermore, to describe in detail the effect of some therapeutic interventions, such as anti-aggregation drugs, a more detailed model of intracellular aggregate formation will have to be included. To this end, further development of the model will incorporate aggregate removal processes and their role in triggering intracellular aggregation, providing a more detailed link to the molecular-level processes within cells^26^.

In addition to more detailed links to molecular aggregation processes, future extensions of this work will focus on broadening the applicability of our model to other neurodegenerative diseases, such as Alzheimer’s and Parkinson’s disease. It will be particularly valuable to explore whether the disease mechanisms highlighted in progressive supranuclear palsy (PSP) are consistent across other tauopathies and proteinopathies. Such expansions could enhance our understanding of these complex diseases and inform general therapeutic strategies.

### Conclusions

In conclusion, we have developed a framework to derive mechanistic information of human tauopathy from digital pathology data processed to detect aggregated cell and nuclear positions. We applied these to tau aggregated cells in *post mortem* tissue from people with PSP. To demonstrate the mechanistic information of these measures and allow for more detailed analysis of patient data, we use a simulation model of cellular aggregation. Although aggregation appears random at short ranges, a spatial coupling effect is revealed at longer distances, on the order of millimetres. In particular, there is increased propensity to aggregate in dense cell regions at the later stages of PSP, consistent with the cell-to-cell triggering mechanism being dependent on the spatial separation of cells. All these observations are successfully explained by our minimal model in which an aggregated cell can trigger aggregation in a healthy cell, a process that is easier the closer cells are closer together. The fact that this approach works and can predict the aggregated cell patterns observed in human brains is surprising and demonstrates that two simple microscopic mechanisms underpin the aggregation in PSP. Finally, using the mechanisms determined from the data, we showcase in simulations how alterations of specific rates would affect the disease progression. This type of modelling can thus form the basis of prediction of drug efficacy for novel therapies to treat or prevent neurodegeneration.

## Methods

### Image analysis pipeline

We utilised an image analysis pipeline (Fig. S4) to analyze immuno-stained brain images obtained from the Cambridge Brain Bank. We first segmented the grey matter parts of the images, followed by colour deconvolution to separate signals from different targets, which in our case are aggregated cells and cell nuclei. We then identified aggregated cells and cell nuclei by thresholding the colour intensity and removed artefacts. The identified aggregated cells and cell nuclei are characterised by several metrics, such as the size, the eccentricity, and the x,y position in the 2-dimensional plane. All the details of the feature extraction can be found in the work by Pansuwan et al.^22^. The extracted features of the objects are then further processed into nearest neighbour distance distribution (NNDD) plots and rolling density plots.

#### Nearest neighbour analysis

To study the spatial arrangement at short distances, we use nearest neighbour distance distribution (NNDD). This is achieved using the cdist function from Python’s scipy.spatial.distance package to calculate the distance between every possible pair of points from a lost of the positions of aggregated cells or nuclei. After calculating all the distances, we sort them for each point to identify the nearest neighbour distance for each point.

#### NND distribution for a purely cell-autonomous system

The nearest-neighbour distance (NND) distribution describes the probability of finding the nearest aggregated cell at a distance *r* from a reference point, assumed here to be the origin. This is derived in two parts: (1) the probability of no aggregated cells within a radius *r*, and (2) the probability density of finding an aggregated cell in the ring *r* → *r* + *dr*.

**Part 1: No Aggregated cells Within Radius** *r* In the cell-autonomous limit, cell aggregation events are random and independent, following a Poisson distribution. The expected number of events within a circle of radius *r* is *λ* = *πr*^2^*D*, where *D* is the overall aggregated cell density. The probability of finding no aggregated cells within radius *r* is:

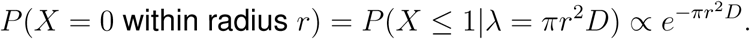

**Part 2: Aggregated cell in the Ring** *r* → *r* + *dr* The probability of finding exactly one aggregated cell in the ring *r* → *r* + *dr* is:

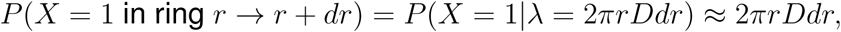

valid for *dr* → 0.

**Final NND Distribution** Combining these, the NND distribution is:

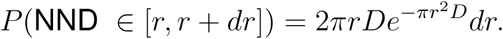

This describes the NND distribution for randomly distributed aggregated cells.

In the opposite limit, when an aggregated cell triggers its direct neighbours and there is negligible cell-autonomous aggregation, then the brain effectively partitions into fully aggregated and non-aggregated regions. The density of aggregated cells in the aggregated regions is simply the density of all cells, so in this limit the NND distribution is again given by the same functional form, 2*πrD e^−πr^*^2^*^Dc^ dr*, except that the relevant density is that of cells, *D* aggregate density as in the cell-autonomous limit. rather than the overall

#### Radial distribution function as a mm-length scale spatial measure

The radial distribution function (RDF) describes how density varies with the radial distance *r* from a reference particle. It can be defined as

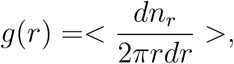

where *dn_r_* is the number of particles within a ring of radius *r*, and width *dr*. We use RDF to characterize the *mm*-length scale aggregated cell and nucleus distributions. We also compute the normalized RDF defined as 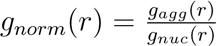, where *g_agg_*(*r*), and *g_nuc_*(*r*) are RDF of aggregated cells and nuclei, respectively. Unlike the more standard definitions of the RDF, this normalised RDF achieves a values of 1 at distances where the system is fully aggregated. Moreover, the normalisation by the RDF of cells also removes contributions to the aggregate spatial patterns from the non-uniform spatial arrangement of cells. This is crucial to obtain RDFs that are interpretable between different brain slices and images which all display different geometries. To perform these calculations, we use the Python package rdf2d, which computes the RDF based on the positions of the aggregated cells or nuclei. This function takes the particle positions and a distance interval (dr=50 µm) to group the distances for analysis, returning two outputs: *g*(*r*), which contains the calculated RDF values, and *r* (radii), which are the distances at which the RDF is evaluated. These results are stored in a dictionary, with both the RDF values and corresponding radii for further analysis or storage.

#### Intra-brain region analysis

##### Preparation of rolling-density data

To quantify the density variation on a length scale larger than the cell level but smaller than a full brain region, we first need to smooth the discrete detection of aggregated cells over space. To compute the local density within a brain slice, we use a rolling density calculation. We divide the brain slices into regions according to a grid and calculate the density of the aggregated cells in each 100 µm by 100 µm region. We select 100 µm as the window size to avoid (i) a too large window size that removes relevant spatial features, and (ii) a too small window size that results in stochastic variation of density. By testing various window sizes (see Fig. S14), we established that a window of 100 µm achieves the best fitting results. Finally, to further smooth the image, we calculate the rolling density. To do so, we select one grid and average the density values of the 7 by 7 grids area that surrounds it. The nucleus and aggregated cell densities for each brain slice are all rolling-average pre-processed and stored for latter usage.

##### Segmentation based on nucleus density

We plot the histogram of the rolling-averaged nucleus densities and then fit the histogram with a Gaussian distribution. We classify the densities into three groups based on the Gaussian distribution: high, medium, and low densities. The boundaries of each group were chosen based on the fitted Gaussian distribution.The first boundary was set to be the *µ* − 2*σ*, where *µ* and *σ* are the mean and standard deviation of the Gaussian distribution, respectively. The data below the first boundary is ignored to exclude the background. The second and third boundaries are set to be the mean plus/minus half of the standard deviation of the Gaussian distribution. The coarse-grained nucleus regions are then used as a mask to separate aggregated cells into different nucleus density regions. This allows us to compare aggregated cell densities in different nucleus density regions, which provides an additional measure to characterise the aggregated cell distribution.

##### APN distribution and APN as a function of nucleus density

Aggregate-per-nucleus (APN) value can capture local aggregation percentage without being affected by variations in cell density. It is calculated by dividing the rolling average density of aggregated cells by that of total cells. One of the important characteristics of APN value is that it can distinguish cell-autonomous and non-cell-autonomous mechanisms when there is variation of cell density in the system. For a cell-autonomous system, no matter how dense the cells are, the aggregated cell number will always be proportional to the total cell number, since the cell-autonomous mechanism, by definition, is independent of cell-to-cell separation. This gives rise to a homogeneous APN value even if cell density varies. By contrast, when there is spatial coupling, high-cell-density regions have larger APN values than low-density regions. Since we can segment the brain slice by the cell density, the APN value can be computed for different cell densities and used as a guide to mechanisms.

#### Pseudo-temporal axis

To understand the temporal evolution of the disease, a temporal dimension for the neuropathological data is needed. However, due to the variability of individuals, a universal time axis across all *post-mortem* data is impossible. Despite such limitations, the well-defined neuropathological staging system can give us an estimation of the disease progression. It was found that the sequential distribution of pathology is associated with the clinical severity in PSP^24^. In addition to this established staging, we also use the fraction of aggregated cells to put different brain regions and individuals on a common axis of pathological severity.

### Model construction

To capture the effect of the cell distribution in each individual, we build a model for protein aggregation in a tissue. Fig. 1 shows the schematics of such a model. Within the cell, *in vitro* protein aggregation mechanisms, including primary nucleation, growth and multiplication, govern the proliferation of protein aggregates. These aggregate formation processes may compete with removal processes. When the balance shifts to net accumulation of aggregates, or if there is a significant seeding event, the cell enters a runaway aggregation state^5,26^. Based on the detailed mathematical treatment in e.g. Thompson et al.^26^ we here coarse-grain this into a switch between a healthy and a runaway aggregation state. Between cells, spatial coupling factors, such as seed transfer or inflammation, allow cells in the runaway aggregation state to exert an *aggregation pressure* on other cells to also switch from the healthy to the runaway aggregation state. In this aspect our model closely mirrors the *Susceptible-Infected-Recovered* (SIR) models of epidemiology^38–40^, although we do not include a *Recovered* state and the probability of “infection” is dependent on the spatial separation, which is fixed. Furthermore, we can assign different vulnerabilities to cells, reflecting the fact that different cell types are present and that each cell may have a different level of resistance against protein aggregation. The following are the detailed explanation of each mechanism we consider.

#### Triggering

Triggering is the switching of a cell from the healthy state to the runaway aggregation state. This switch may for example occur when aggregate production outweighs removal^26^, or in the case when removal processes are negligible, when the first self-replicating aggregate appears either by nucleation or seeding^5,41^. Our model is agnostic to the type of trigger and includes it as a stochastic process.

#### Spatial coupling factors

There is ample experimental evidence that pathology and, at least in model systems, also aggregated species can be transferred from one cell to another, for example along axonal connections, but potentially also through extracellular space^42,43^. In order to model this potential of cells in the runaway aggregating state to trigger healthy cells in a general manner, we define an *aggregation pressure*. This is used to compute the probability to trigger a healthy cell and depends on the relative positions of the cells involved. Between brain regions, information on the connectivity exists, but on the length scales studied in this work, a determination of the connectivity of each individual cell is far out of reach of current experimental techniques. Thus, we define an aggregation pressure that depends only on the spatial separation of two cells. The overall aggregation pressure on a given healthy cell is then computed by considering the aggregation pressures of all aggregated cells in its vicinity.

For the majority of this work, we assume an aggregation pressure that is Gaussian in the distance between the aggregated and the healthy cell. The aggregation pressure on a cell *i* in each time step Δ*t* is then defined as *p_s_* = 1 − *e^−λi^*, where 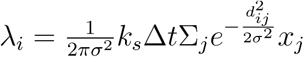 is the average aggregation events per time step Δ*t*, where *σ* is the Gaussian diffusion radius, *d_ij_* is the pairwise distance between cell *i* and *j*, *x_j_* is the value quantifying whether cell *j* is aggregated (*x_j_* = 1) or not (*x_j_* = 0), and *k_s_* is the rate of cell-to-cell triggering. To ensure our conclusions are general, we also show that changing the functional form of this effect does not change our conclusions, see *Modelling cell-level aggregate formation*.

#### Selective vulnerability

Our model also incorporates the selective vulnerability of each cell. Though the definition of selective vulnerability varies in different contexts^44^, here we define the selective vulnerability as the cell’s ability to resist the switch to a runaway aggregation state, by both cell-autonomous or external triggers (Fig. 1A). Mathematically, we define the vulnerability constant *v_i_* as a value between 0 and 1 for each cell *i*, which simply multiplies the probability of triggering to produce an updated probability that takes into account the vulnerability. In future, it may be possible to estimate the values of *v_i_*, for example through spatial transcriptomics coupled with a detailed understanding of which genes are govern vulnerability. As such data are however not yet available, in our model, we consider a number of different *v_i_* distributions, such as *v_i_*= *const.* or *v_i_*= *U* (0, 1), where *U* (0, 1) is the uniform distribution between 0 and 1.

### Simulation parameters

The simulation parameters for Fig. 2, S8, & S9 are as follows. We simulate a box containing 10,002 cells. The code generates a random spatial distribution of cells while controlling the average cell-to-cell distance, which is set to 27.3 µm, a value chosen based on measurements from the data. A higher density (or smaller average distances) is imposed in the middle third of the y-axis range, with an average cell-to-cell distance 0.8 times shorter than in the other sections. The x-coordinates are randomly and uniformly distributed across the entire x-axis range while the y-coordinates are adjusted randomly within each density band.

Mechanistic parameters are chosen to represent different spatial coupling conditions. For no spatial coupling, *k_s_/k_a_*= 0.001 is used. For short-range and long-range spatial coupling, *k_s_/k_a_*= 10000 is applied with *σ* = 40 µm for short-range coupling and 400 µm for long-range coupling. Periodic boundary conditions are assumed for the simulation.

### Parameter inference

Our computational model not only identifies the dominant molecular mechanisms but also allows us to quantify their rate constants using simulation based inference. To do so, we reconstruct a virtual brain slice *in silico* based on the cell positions from a histopathological image and then compare simulations on this cell arrangement with experimental data using various spatial measures from the data and the simulation. The unknown parameters to be determined are the rate of cell-autonomous triggering (*k_a_*), rate of cell-to-cell triggering (*k_s_*) and the cell-to-cell coupling radius (*σ*).

Because our data is an endpoint measure rather than a time-course, we can only determine the ratio of rate constants, not their absolute values. We are thus left with parameters *k_s_/k_a_* and *σ* to be fit. To perform the fits, we run simulations with a combination of parameters {*k_s_/k_a_* ∈ [0.001, 0.001, 0.01, 0.1, 1, 10, 100, 1000, 10000, 100000]} ⊗ {*σ* (*µm*) ∈ [50, 100, 200, 400, 600, 800]}. At each set of parameters we perform 10 repeats of the simulation, to account for stochasticity. To find the best fit of *k_s_/k_a_*, we compare the histogram of APN values (all the histograms discussed are normalised to 1) from the data and the simulation. We define the error of each repeat of the parameter set as the mean of bin-by-bin squared difference between the APN histogram of this repeat and the data. We then compute the mean error across 10 repeats. The two-dimensional plots of the mean error across different {*k_s_/k_a_*} ⊗ {*σ*} are shown in Fig. S10. Additionally, we used the nearest neighbour distribution to establish a lower bound for the spatial coupling radius *σ* (Fig. S11).

Although other readouts can be compared, such as RDF (see Fig. S12), we select APN histogram and nearest neighbour distribution as our readouts for fitting because of their richness in mechanistic information.

Given the lack of independent data on vulnerability to aggregation, we assume a constant vulnerability in this analysis. However, the vulnerability and cell-to-cell coupling radius are coupled, leading to some uncertainty in the coupling radius due to the uncertainty in vulnerability. More specifically, a system with strongly varying vulnerability and a short coupling radius and a system with a weakly varying vulnerability and a large coupling radius give rise to similar patterns of aggregation, see Fig. S9. The assumption of a constant vulnerability for all cells thus produces an upper bound estimate for the coupling distance.

### Derivation of the switch fraction

Let *f* (*t*) be the fraction of aggregated cells. The rate at which *f* (*t*) is increased due to cell autonomous processes is simply proportional to the fraction of unaggregated cells, thus

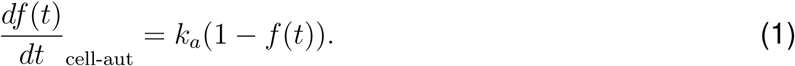

The contribution from cell-to-cell triggering is significantly more complex as it depends on the specific patterns of aggregated cells. However, for a spatially uniform system it can be assumed to be proportional to both the fraction of aggregated cells and the fraction of unaggregated cells, giving

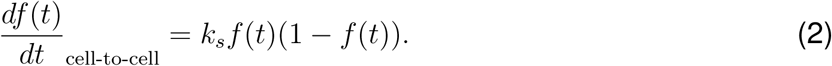

Note the similarity here to the auto-catalytic amplification term in a Fisher-KPP equation, as used in^10^. The two rates are equal at the switch fraction *f* (*t*) = *f_s_* thus *k_s_f_s_* = *k_a_* giving an estimate for the switch fraction simply as the ratio of the rate constants as quoted in the main text.

## Data Availability

All data produced in the present study are available upon reasonable request to the authors.

## Ethical approval

Human post-mortem brain tissue was acquired from the Cambridge Brain Bank (Cambridge University Hospitals). The Cambridge Brain Bank is supported by the NIHR Cambridge Biomedical Research Centre (NIHR203312). We gratefully acknowledge the participation of all our patient and control volunteers.

## Supplementary Material

### Inter-brain-level analysis of PSP brains

The APN value per brain region is calculated for each patient, categorised by Kovacs stages^24^. The relative aggregate amounts in different regions are approximately maintained across stages (Fig. S1A). A logarithmic plot of the APN value (Fig. S1 B-M) reveals consistent increase across brain regions. However, the real temporal interval between stages has not yet been determined. It is known that gliosis may lead to the increase of cell density with the progression of neurode-generative diseases^45^. We observed similar trends of cell density increase with disease stage in our data. The mean cell densities increase from ∼ 1000*/mm*^2^ to ∼ 1400 */mm*^2^, a 1.4-fold increase, from stage 2 to stage 6 (Fig. S2). This increase fold also matched the value in AD (about 1.2 fold increase)^45^. Per-brain-region analysis also shows there is a correlation between nucleus density and stage in most brain regions (PMC, S1, OC, STR, M1, ACC, PC), while other brain regions (STN, GP, CMB) show no correlation or negative correlation (Fig. S2). Investigation of the effect on different cell-types shows that it is predominantly oligodendrocytes that contribute to the growth of the aggregated cell percentage, whereas neuronal and astroglial aggregates show only a mild increase with stage (Fig. S3A-L). This finding suggests that it is oligodendroglial cells that shape the overall evolution of pathology in the disease, which is also consistent with the recent genome wide study^46^.

**Figure S1:**
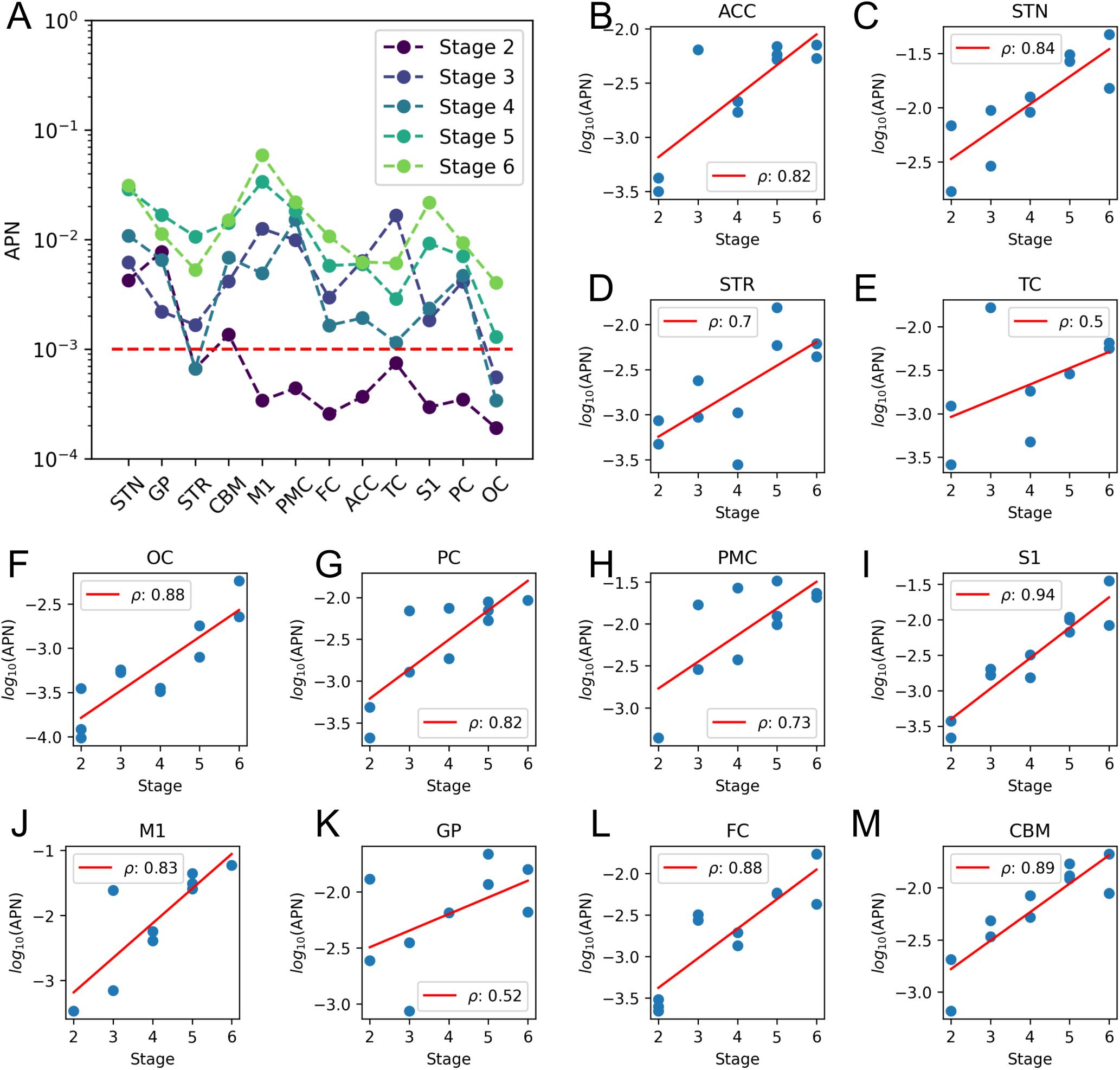
Inter-brain-level analysis reveals aggregated cells increase with stage. (A) The fraction aggregated per brain region in different disease stages. The red line is the switch fraction calculated from *k_s_/k_a_* = 1000 (B-M) Fraction aggregated over staged for different brain regions. *ρ* represents the Pearson correlation coefficient.

**Table S1:** Summary of patient information.

| Patient ID | Sex | Stage | Brain regions |
| --- | --- | --- | --- |
| 1 | Male | 2 | FC,OC,CBM,S1,M1,PMC,PC,TC,ACC,STN,GP,STR |
| 2 | Female | 2 | FC,OC,CBM,S1,PMC,PC,TC,ACC,STN,GP,STR |
| 3 | Male | 3 | FC,OC,CBM,S1,M1,PMC,PC,STN,GP,STR |
| 4 | Female | 3 | FC,OC,CBM,S1,M1,PMC,PC,TC,ACC,STN,GP,STR |
| 5 | Male | 4 | FC,OC,CBM,S1,M1,PMC,PC,TC,ACC,STN,GP,STR |
| 6 | Male | 4 | FC,OC,CBM,S1,M1,PMC,PC,TC,ACC,STN,STR |
| 7 | Female | 5 | FC,CBM,S1,M1,PMC,PC,ACC,STN,GP,STR |
| 8 | Female | 5 | FC,OC,CBM,S1,M1,PMC,PC,TC,ACC,STN,GP,STR |
| 9 | Male | 5 | OC,CBM,S1,M1,PMC,PC,ACC |
| 10 | Male | 6 | FC,OC,CBM,S1,M1,PMC,PC,TC,ACC,STN,GP,STR |
| 11 | Female | 6 | FC,OC,CBM,S1,PMC,TC,ACC,STN,GP,STR |

**Table S2:** List of abbreviations.

|  |  |
| --- | --- |
| FC | Frontal cortex |
| OC | Occipital cortex |
| CBM | Cerebellum |
| S1 | Primary somatosensory cortex |
| M1 | Primary motor cortex |
| PMC | Premotor cortex |
| PC | Parietal cortex |
| TC | Temporal cortex |
| ACC | Anterior cingulate cortex |
| STN | Substantia nigra |
| GP | Globus pallidus |
| STR | Striatum |
| PSP | Progressive supranuclear palsy |
| AD | Alzheimer's disease |
| NFT | Neurofibrillary tangle |
| CB | Coiled body |
| TA | Tufted astrocyte |

**Figure S2:**
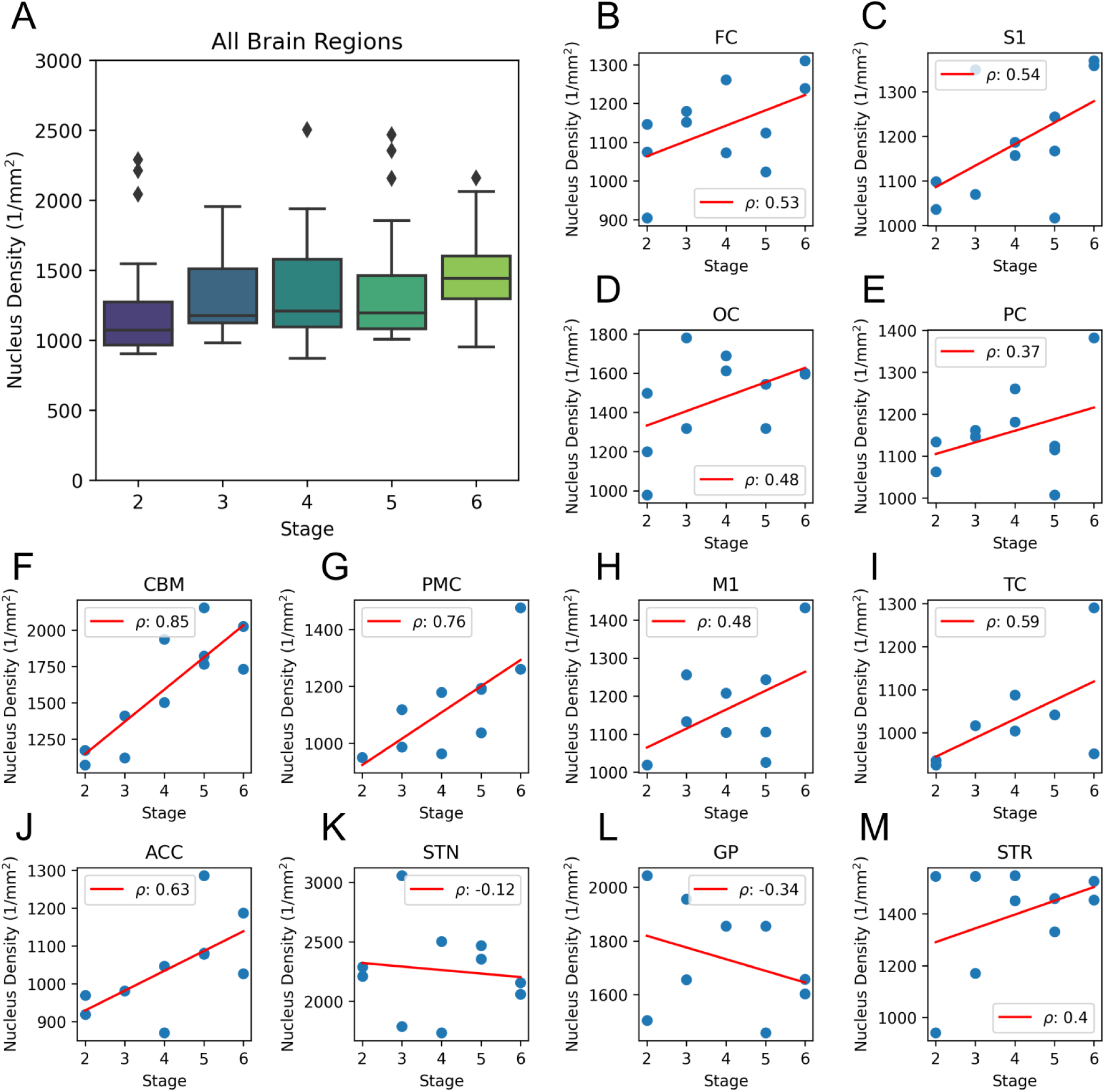
Densities of cell with stage in different brain regions. (A) The fraction aggregated per brain region in different disease stages. The box represents the interquartile range (IQR), encompassing the middle 50% of the data with edges at the first and third quartiles. Whiskers extend to 1.5 times the IQR from the quartiles to show the data range, while points outside these whiskers are plotted as outliers. (B-M) Nucleus density over stage of different brain regions. *ρ* represents the Pearson correlation coefficient.

**Figure S3:**
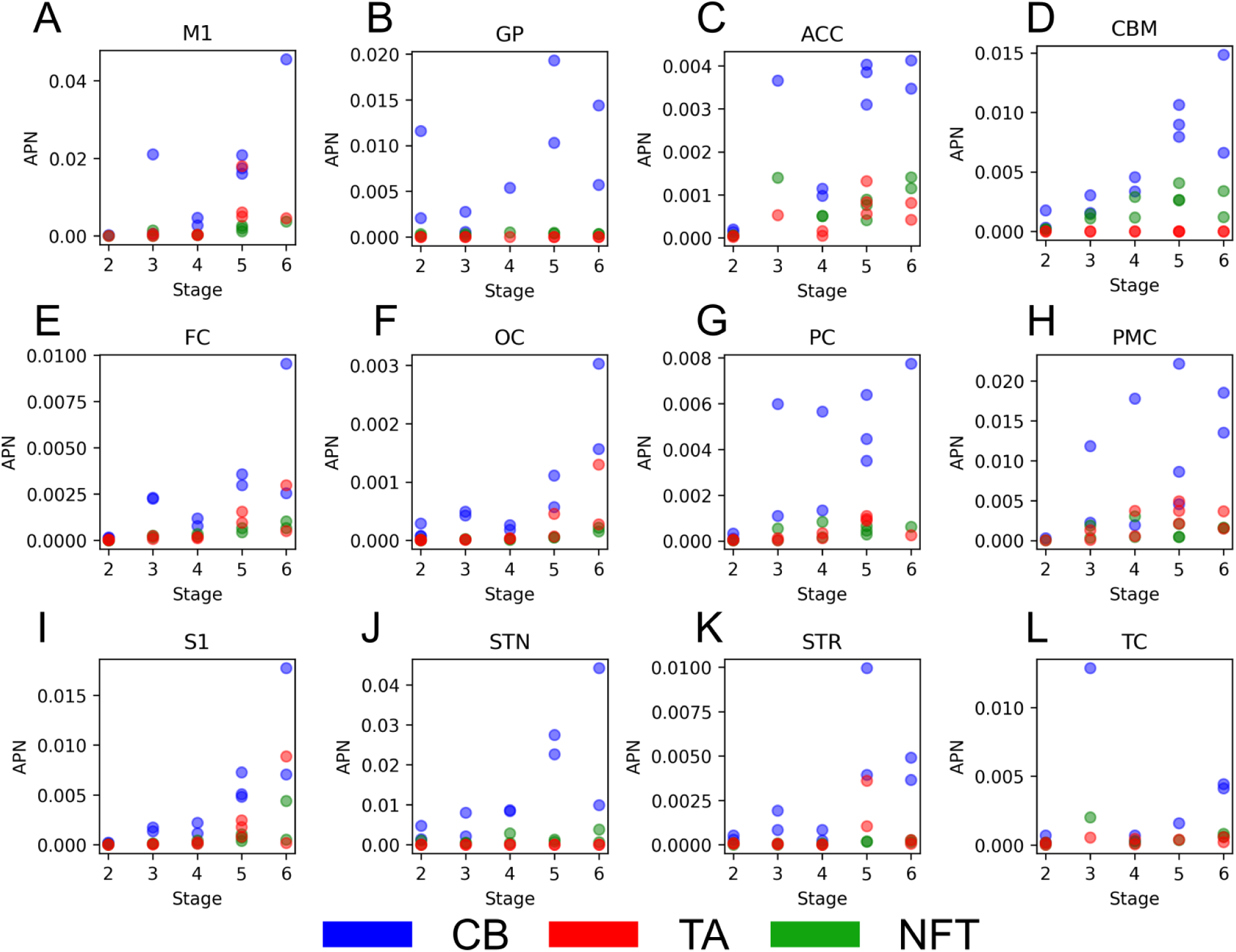
APN values for different cell types across disease stages. Coiled Bodies (CBs), which form in oligodendrocytes, are shown in blue, Tufted Astrocytes (TAs), which form in astrocytes, are shown in red and Neurofibrillary Tangles (NFTs) which form in neurons are shown in green.

**Figure S4:**
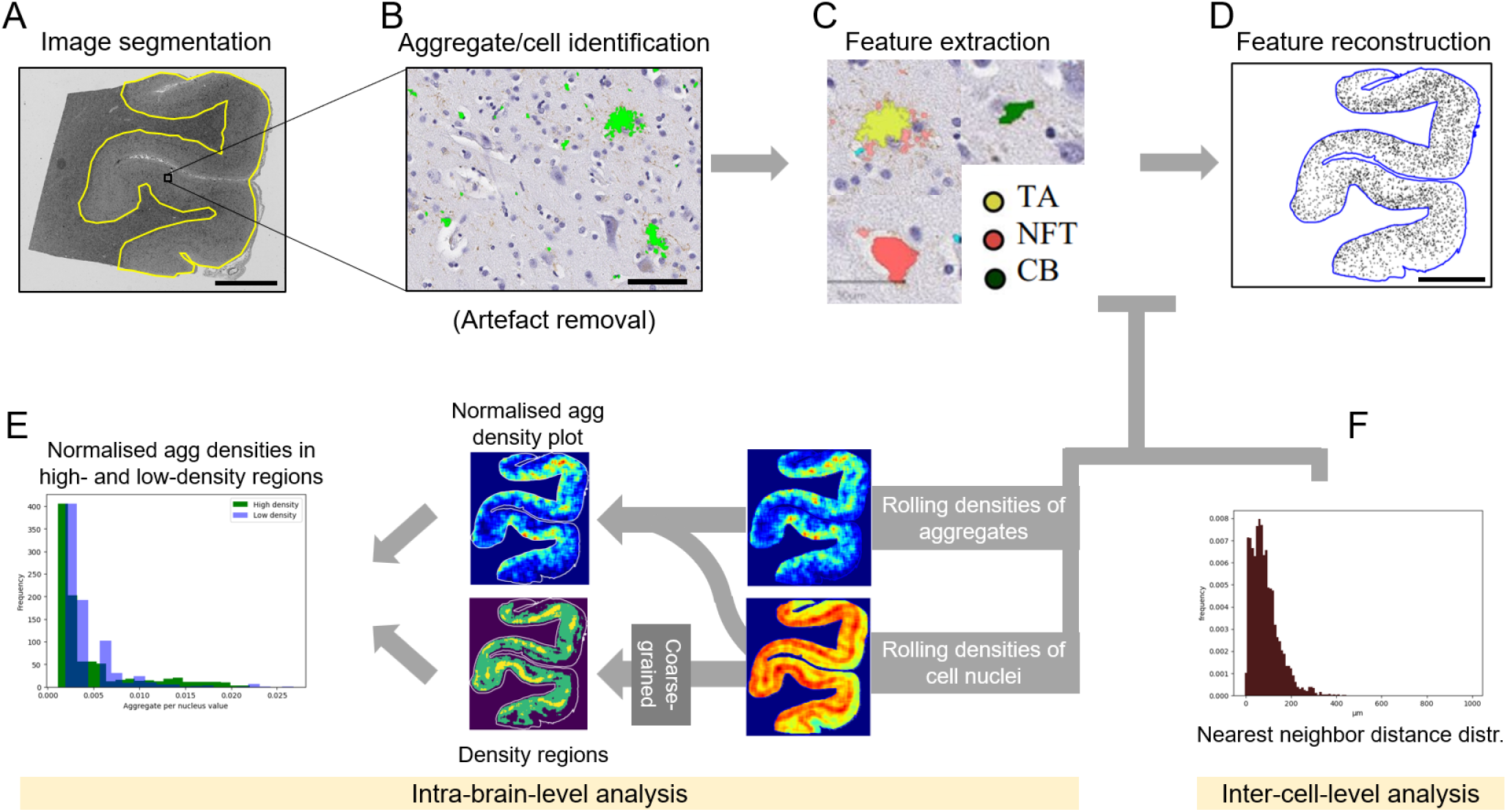
Image analysis pipeline of brain slices. (A) The region of grey matter is segmented (Scale bar = 5 mm). (B) Thresholding and shape classifiers are applied to identify aggregates and nuclei (Scale bar = 50µm). (C) Features of aggregated cells and nuclei, such as size and the spatial locations, are extracted. (D) Finally, a feature-tailored image can be reconstructed (Scale bar = 5 mm). (E & F) Aggregated cell/nucleus features can be further analysed: nearest neighbour distance distribution and rolling density plots can characterize aggregated cell/nucleus patterns on different length scales.

**Figure S5:**
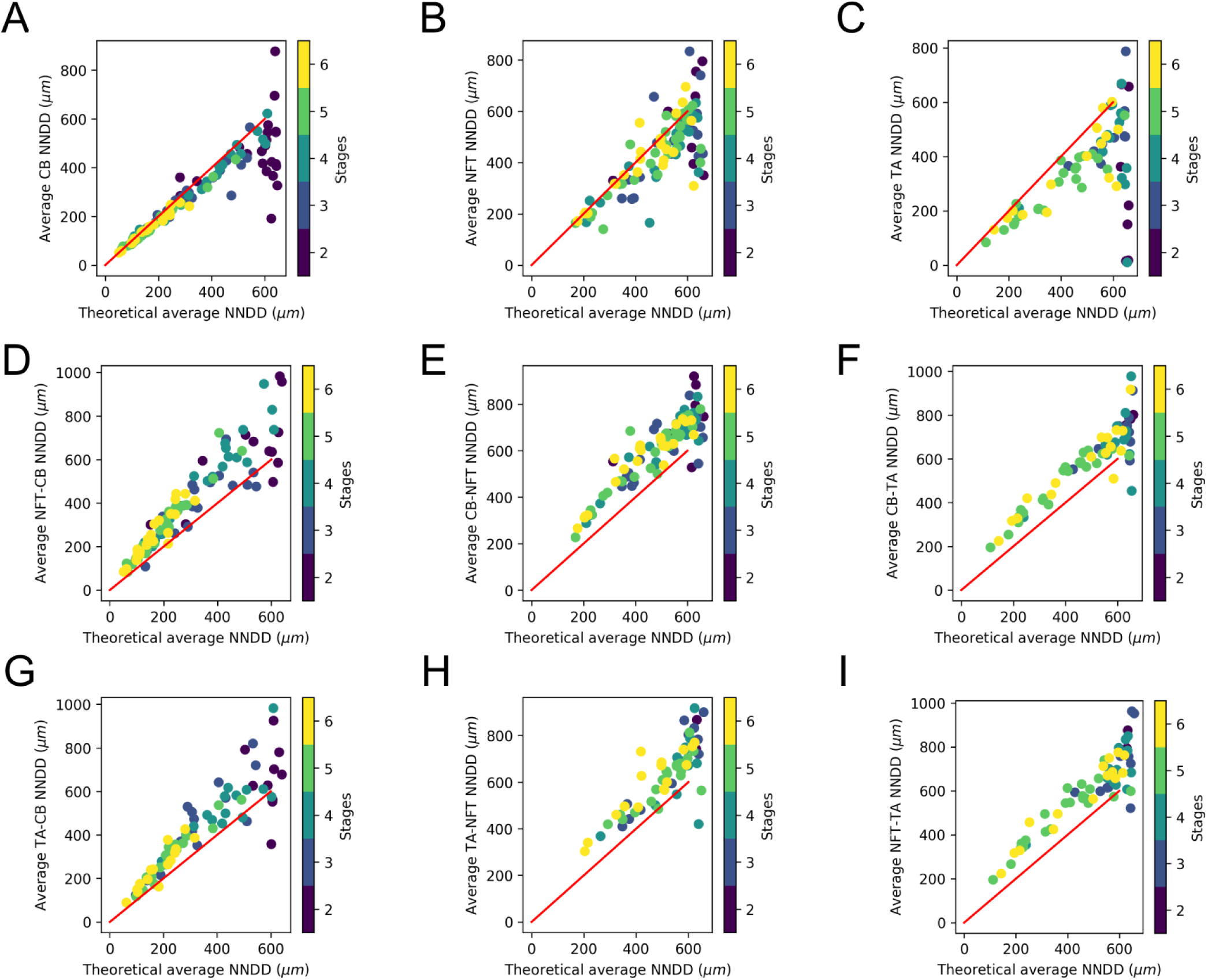
NND distribution within and across different aggregated cell subtypes. The theoretical average NNDD is plotted against the value determined for each brain slice. The theoretical average NNDD for A-C is calculated assuming a random distribution of that particular aggregated cell subtype. The theoretical average NNDD for D-I is calculated assuming a random distribution of the latter aggregated cell subtype. For example, the NFT-CB cross-type NNDD uses the theoretical random distribution of CB.

**Figure S6:**
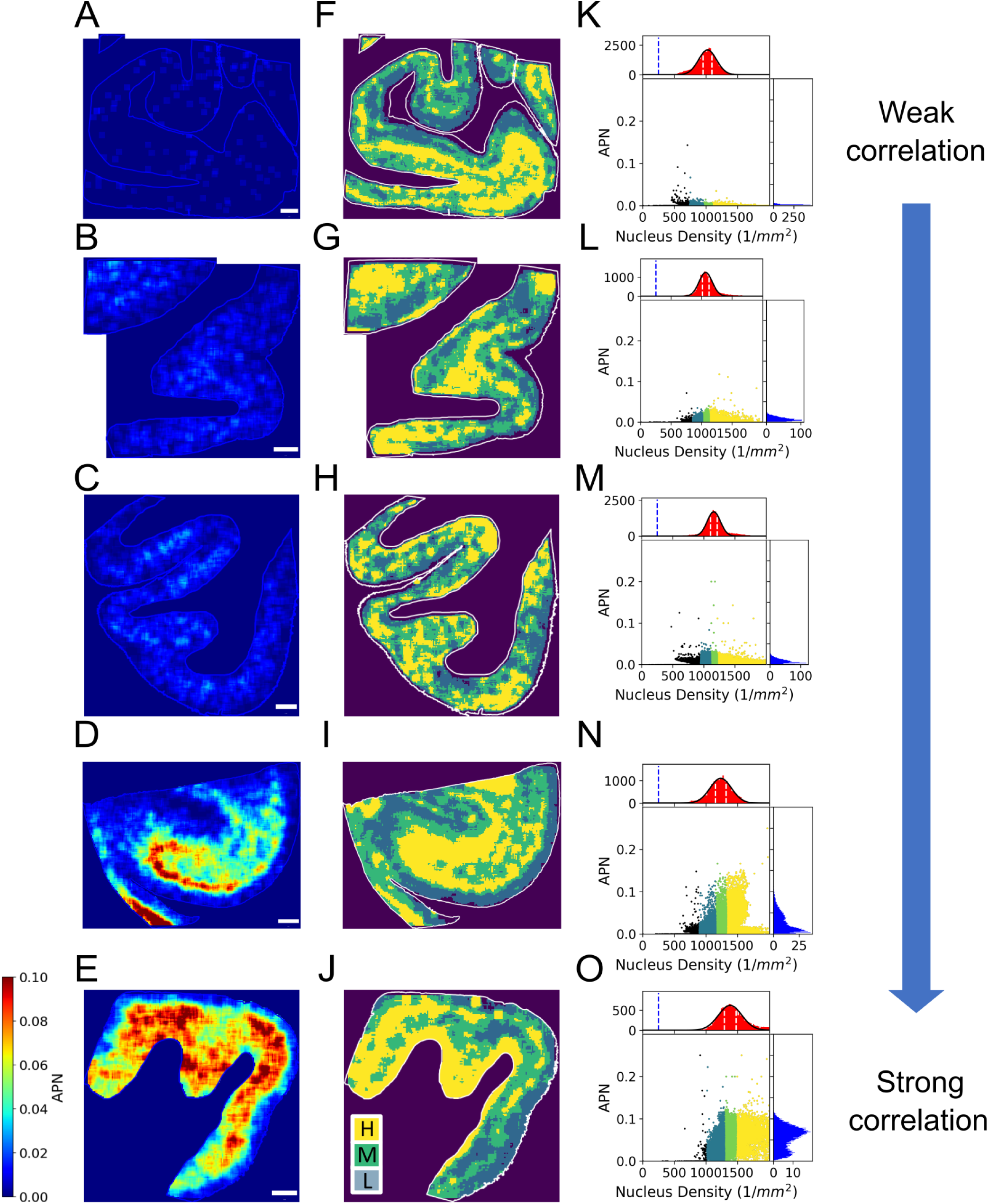
Correlation between APN values and nucleus density with disease stage. (A-E) APN plots for example brain images from Kovacs stage 2 (A), stage 3 (B), stage 4 (C), stage 5 (D) and stage 6 (E). Scale bar = 2 mm. (F-J) Corresponding nucleus density regions of (A-E). High density region: yellow; moderate density region:green; low density region: cyan. (K-O) Pixel-wise correlation plots between (A-E) and (F-J) and their corresponding histograms in two axes. Different colours show different nucleus density regions.

**Figure S7:**
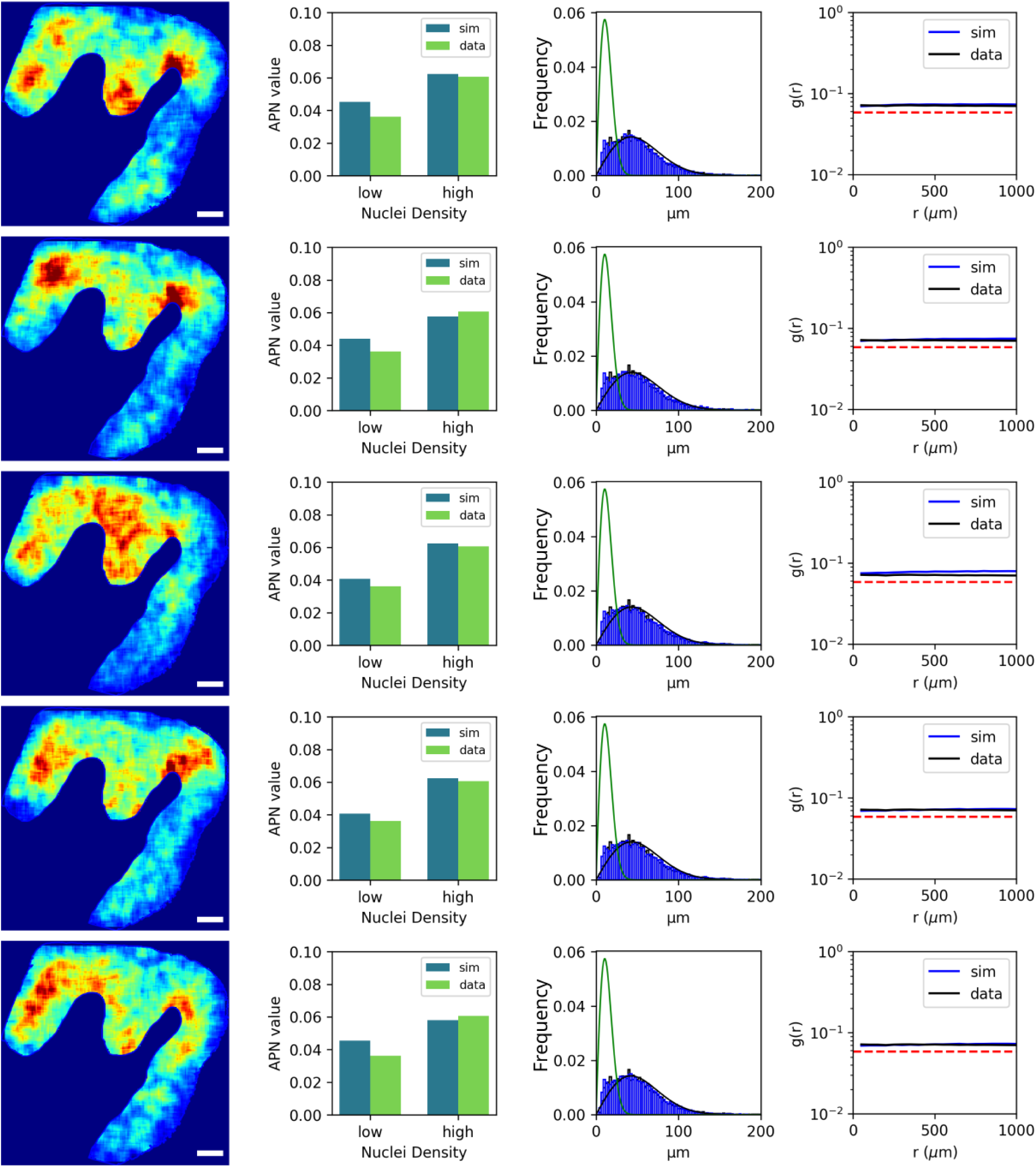
Comparison of repeats of an example simulation. Each row represents one realization of the simulation with the same parameter set. Each row contains the following: 2D patterns of the aggregated cell rolling-average density (far left), APN values in different density regions (middle left), NND distributions (middle right), and RDF (far right).

**Figure S8:**
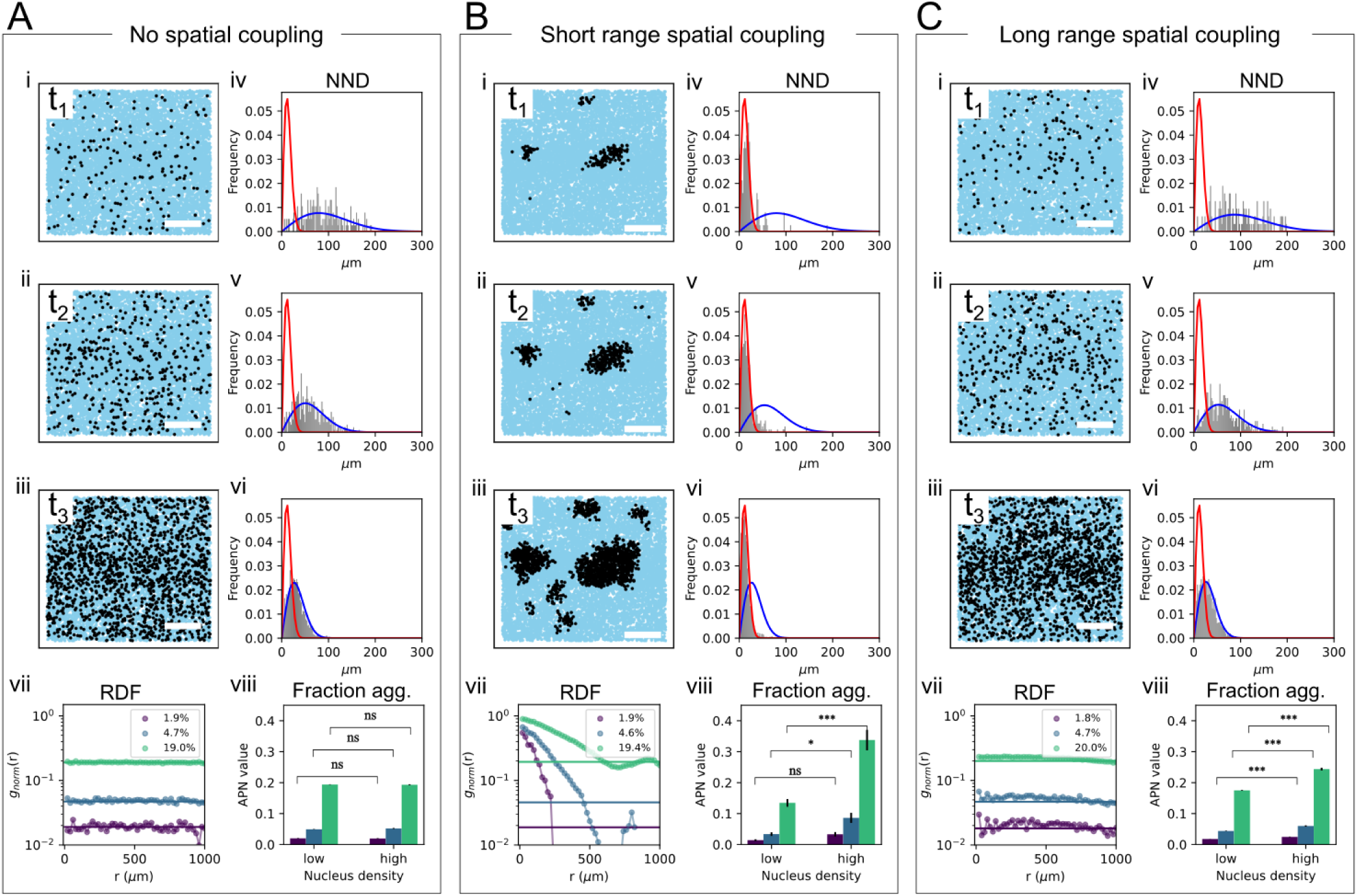
Simulation of aggregation dynamics with exponential decay spatial coupling dependence. The simulation conditions are the same as in Fig. 2 except that the spatial coupling has distance dependence has been changed to *e^−d/σ^*. Thus the spatial coupling strength at a distance of 0 matches between the normal distribution used in the main text, see e.g. Fig. 2) and the exponential decay used here. Scale bar on panels i-iii: 500 µm. The simulation parameters for this figure are provided in Sec. *Simulation parameters*.

**Figure S9:**
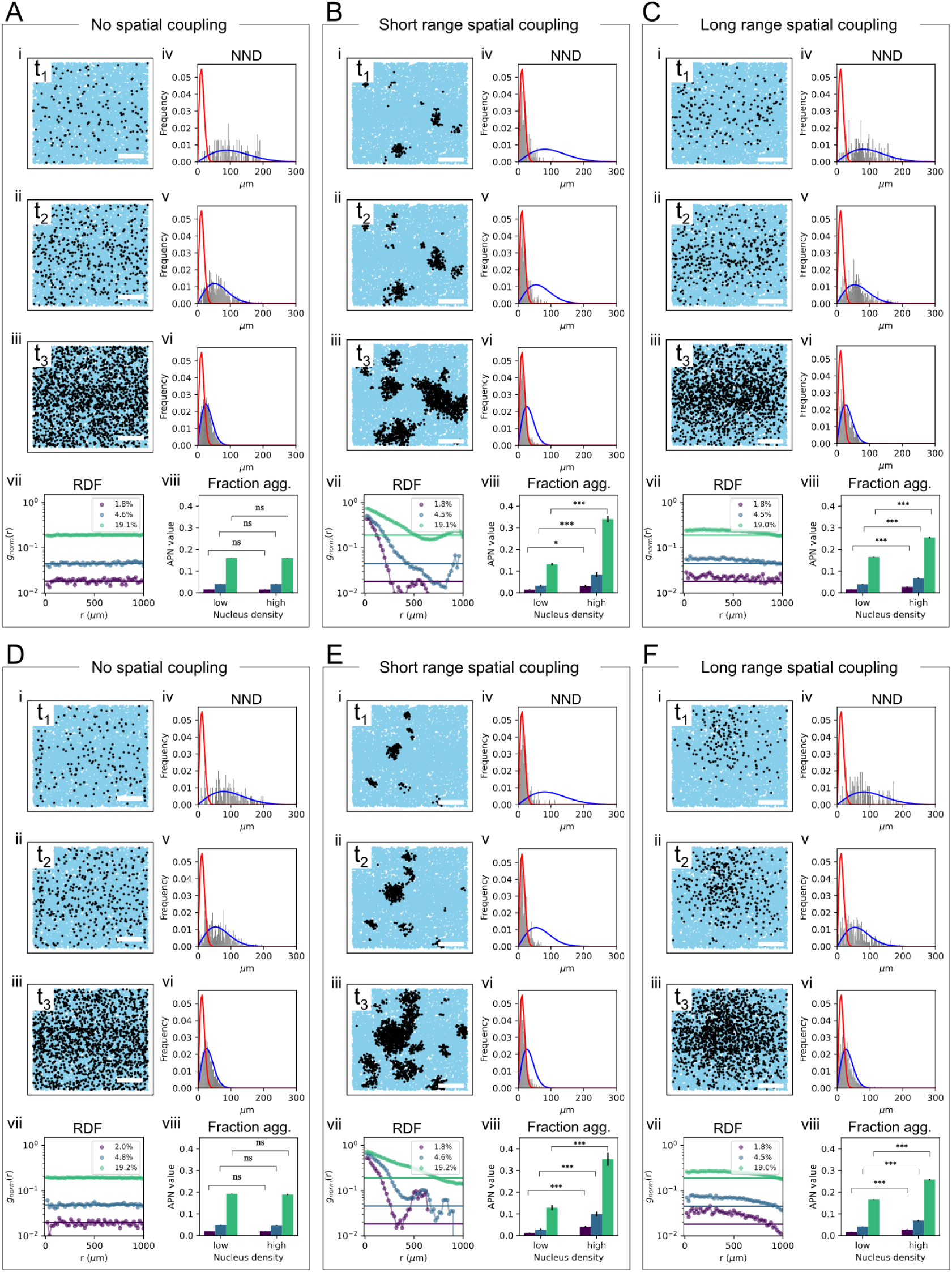
Simulation of aggregation dynamics for Bernoulli and uniform distributions of vulnerability. The simulation conditions are the same as in Fig. 2 except for vulnerability values. (A-C) There are two groups of cells with distinct vulnerability values: one group with a value of 0.1, comprising 95% of the population, and another group with a value of 1, comprising 5% of the population. (D-E) the vulnerability distribution is uniform on [0, 1], *U* (0, 1). Scale bar on panels i-iii: 500 µm. The simulation parameters for this figure are provided in Sec. *Simulation parameters*.

**Figure S10:**
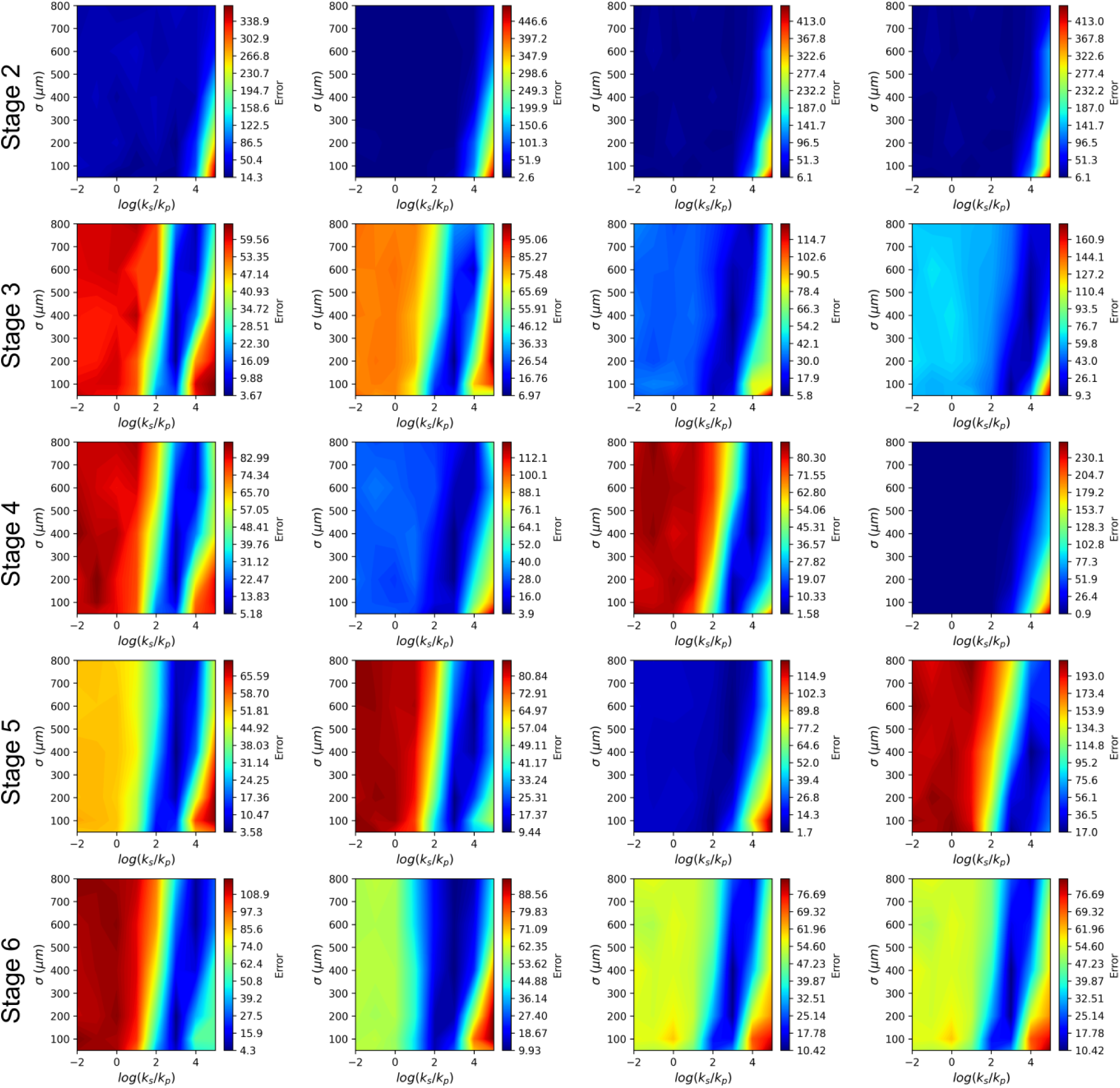
Parameter inference through APN histograms. Each panel shows the two-dimensional plots of the mean error across different parameter sets (see *Parameter inference* for the definition of error). The figure shows the analysis of 20 brain slices arranged in 5 stages, each with 4 panels per stage. Each column represents a distinct brain region: from left to right PMC, M1, PC, and S1.

**Figure S11:**
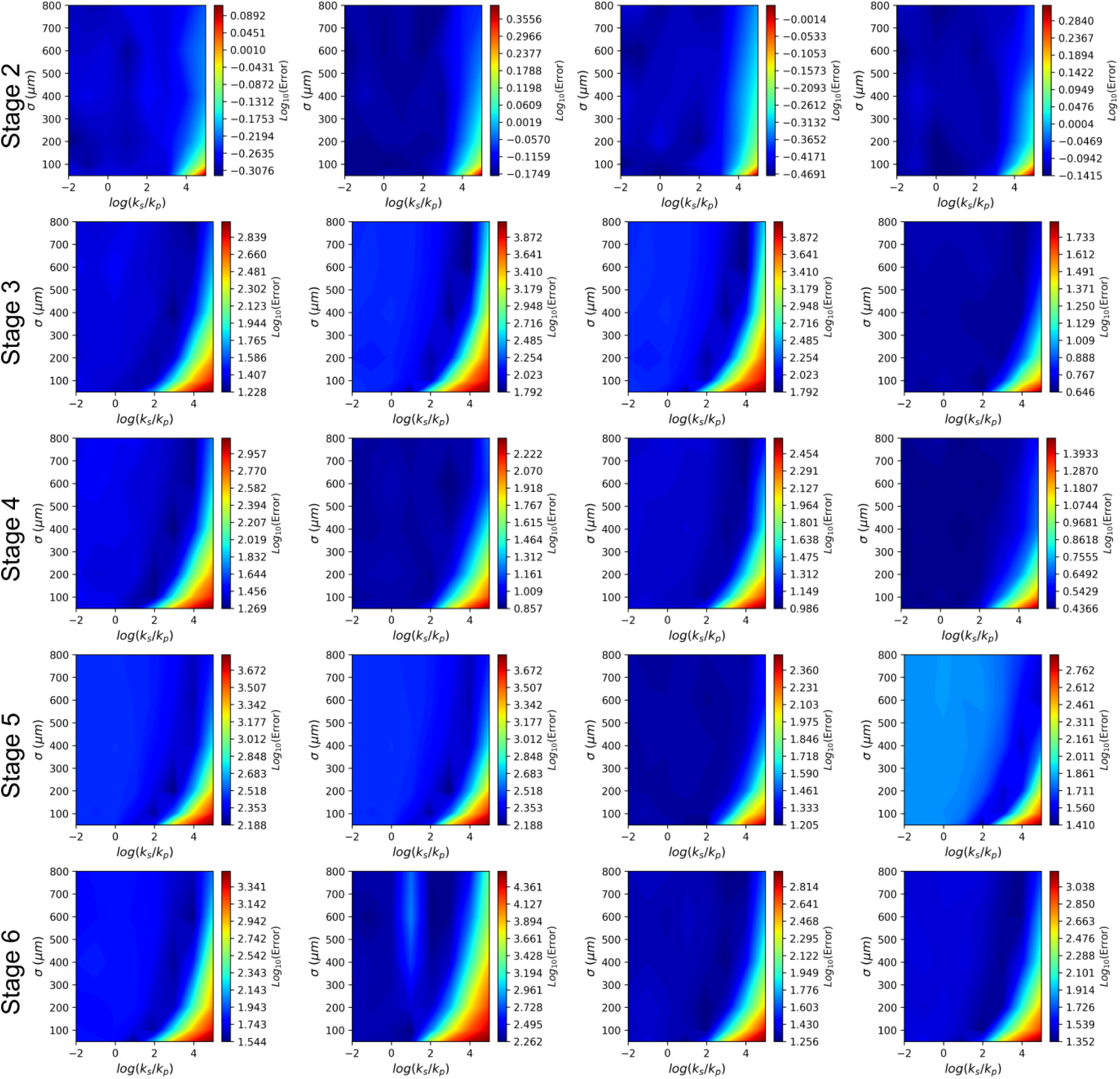
Inference through NNDD. Each panel shows the two-dimensional plots of the mean error across different parameter sets (see *Parameter inference* for the definition of error). The figure shows the analysis of 20 brain slices arranged in 5 stages, each with 4 panels per stage (corresponding to the same brain slices as in Fig. S10). Each column represents a distinct brain region: from left to right PMC, M1, PC, and S1.

**Figure S12:**
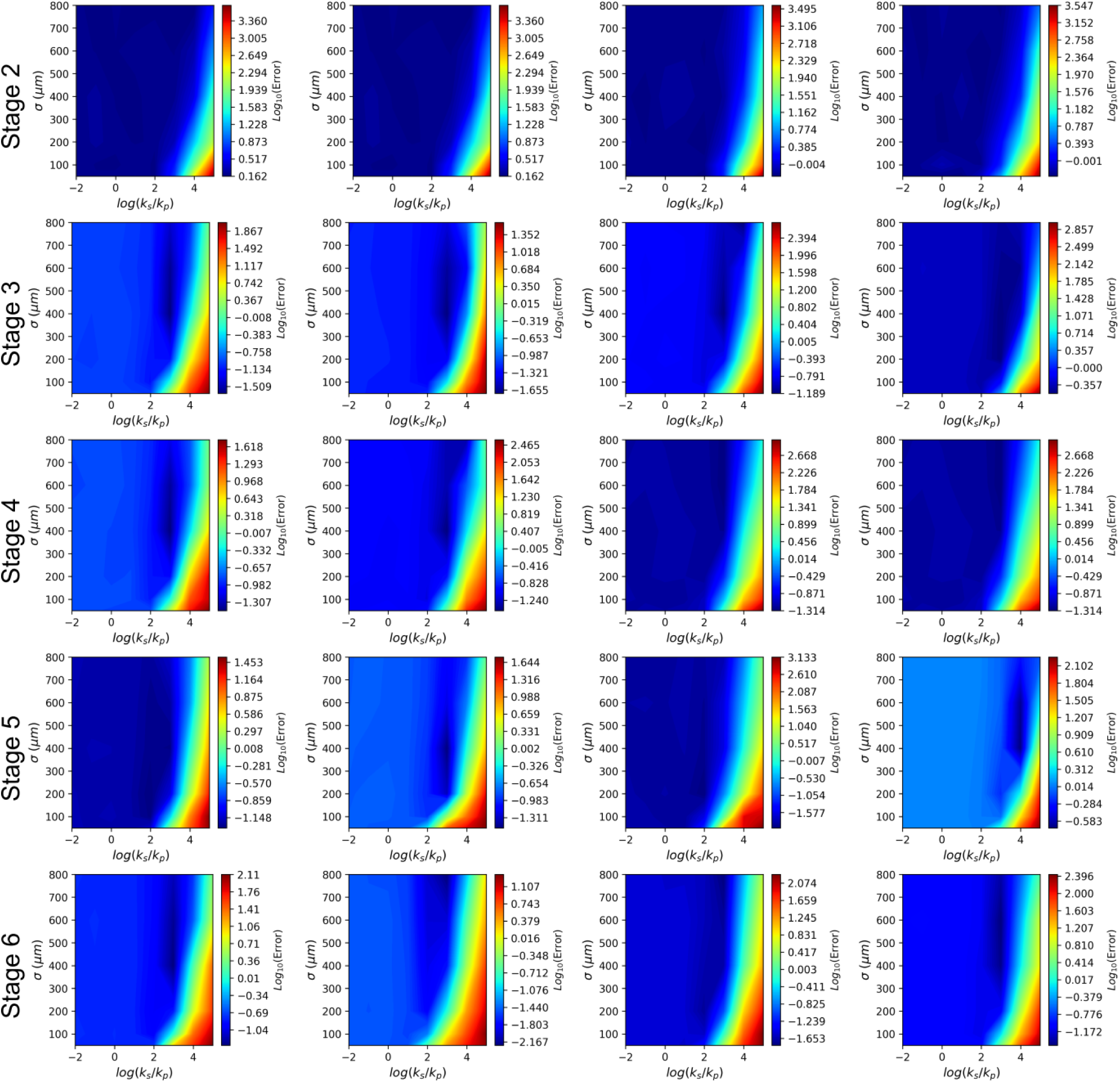
Inference through RDF. Each panel shows the two-dimensional plots of the mean error across different parameter sets (see *Parameter inference* for the definition of error). The figure shows the analysis of 20 brain slices arranged in 5 stages, each with 4 panels per stage (corresponding to the same brain slices as in Fig. S10). Each column represents a distinct brain region: from left to right PMC, M1, PC, and S1.

**Figure S13:**
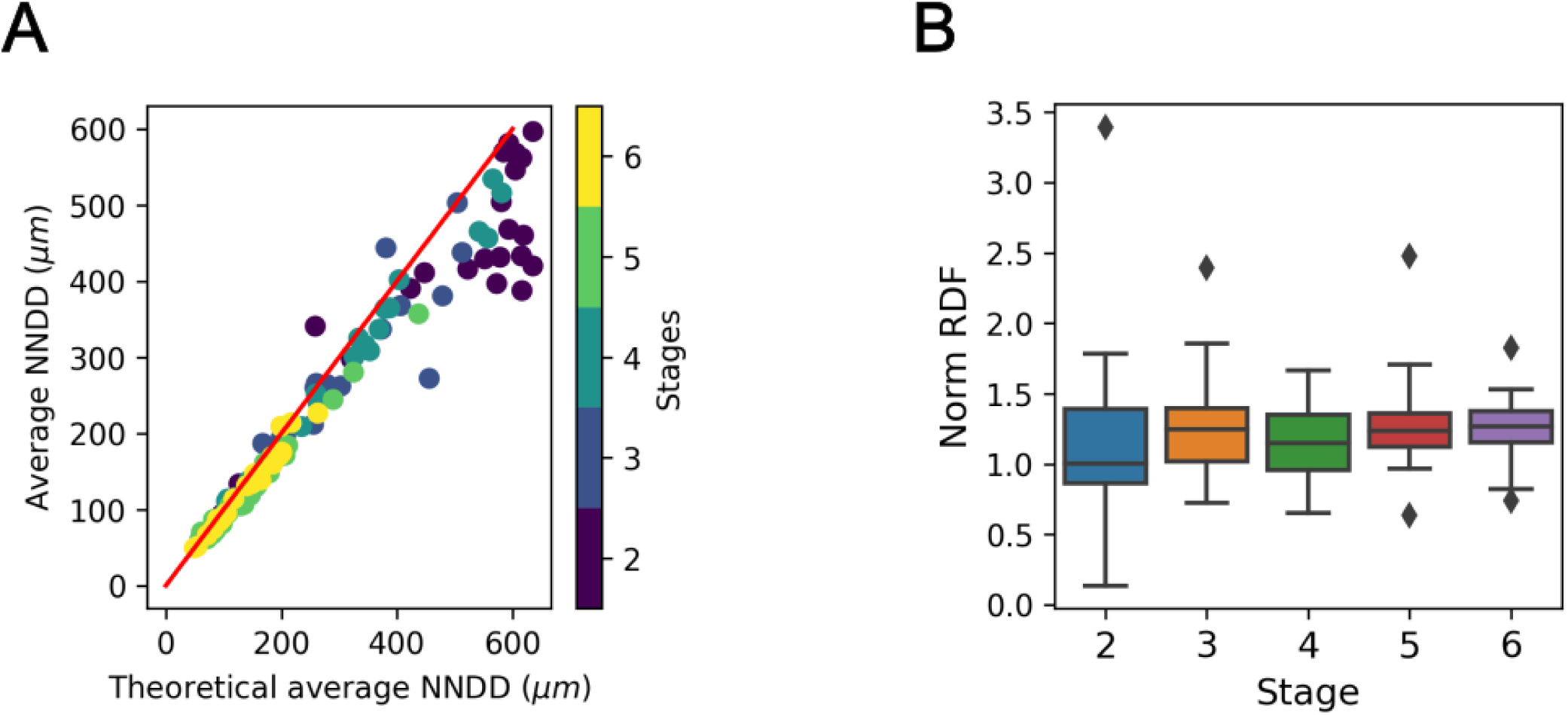
More model-free analysis from the patient data. (A) Average aggregate nearest neighbour distance determined in patient data, compared to that of a random distribution (*R*^2^ = 0.99). The red curve represents the scenario where the measured values match the theoretical values. (B) Box plots showing the ratio between average value of the radial distribution function up to a distance of 1 mm and the average value of hypothetical radial distribution function up to a distance of 1 mm when aggregated cells are randomly distributed, grouped by stage. Values of the average RDF above 1 denote an increased clustering of aggregated cells within clusters of approximately 1 mm. This can be observed at all stages from stage 3 onward. In earlier stages, the number of aggregated cells is too low to draw clear conclusions. The box represents the interquartile range (IQR), encompassing the middle 50% of the data with edges at the first and third quartiles. Whiskers extend to 1.5 times the IQR from the quartiles to show the data range, while points outside these whiskers are plotted as outliers.

**Figure S14:**
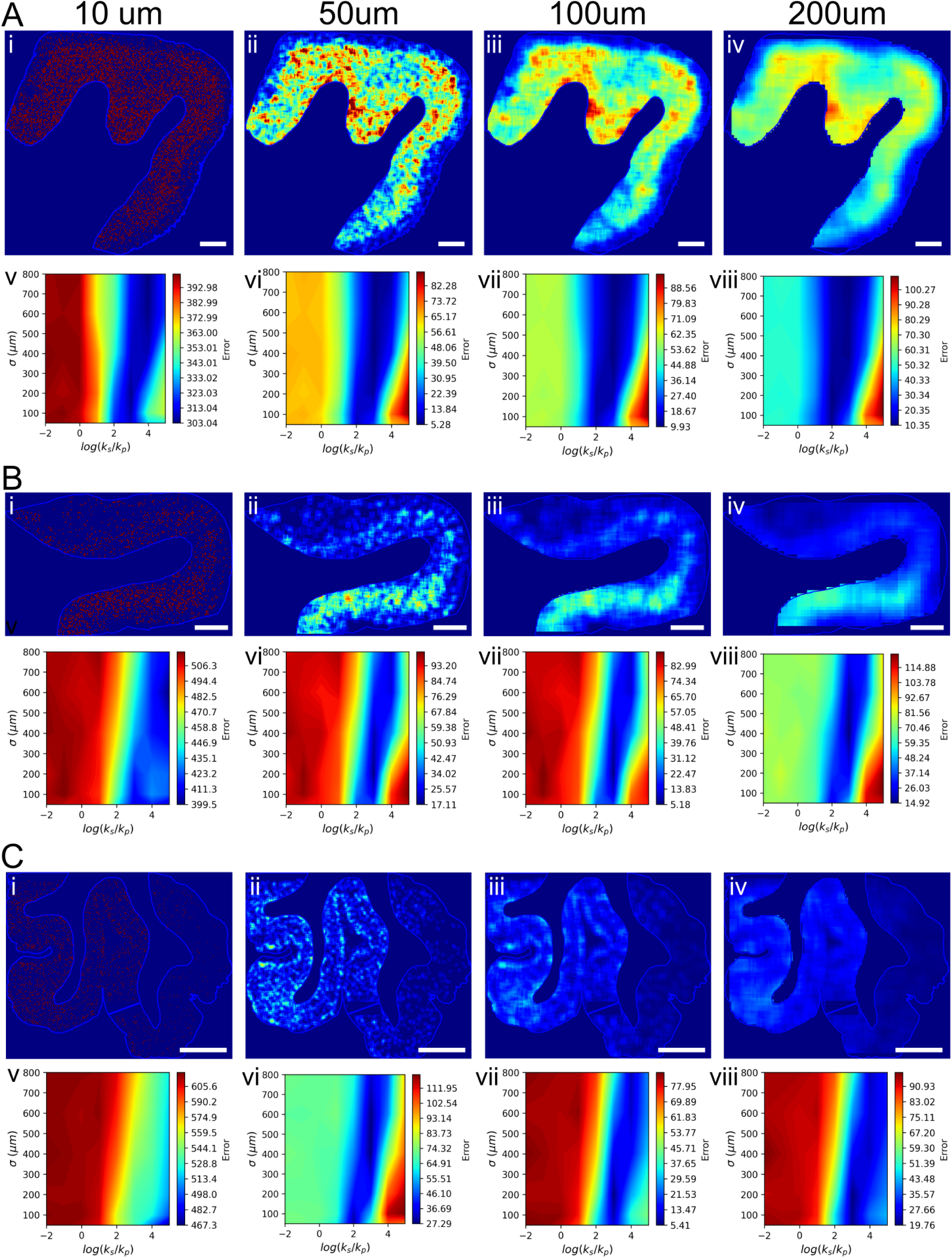
Comparison of different sizes of rolling density window. (A)-(C) Varying rolling density window size from 10 *µm* to 200 *µm* from three example brain slices. In each panel, the top sub-panels show images of different window sizes. The bottom sub-panels shows the two-dimensional plots of the mean error across different parameter sets (see *Methods*) for the definition of error. Scale bar = 2 mm.

## References

1. Chiti, F., and Dobson, C. M. (2006). Protein misfolding, functional amyloid, and human disease. Annual Review of Biochemistry 75, 333–366. URL: https://doi.org/10.1146/annurev.biochem.75.101304.123901. doi:10.1146/annurev.biochem.75.101304.123901. arXiv:https://doi.org/10.1146/annurev.biochem.75.101304.123901. PMID: 16756495.

2. Chiti, F., and Dobson, C. M. (2017). Protein misfolding, amyloid formation, and human disease: A summary of progress over the last decade. Annual Review of Biochemistry 86, 27–68. URL: https://doi.org/10.1146/annurev-biochem-061516-045115. doi:10.1146/annurev-biochem-061516-045115. arXiv:https://doi.org/10.1146/annurev-biochem-061516-045115.PMID: 28498720.

3. Fitzpatrick, A. W. P., Falcon, B., He, S., Murzin, A. G., Murshudov, G., Garringer, H. J., Crowther, R. A., Ghetti, B., Goedert, M., and Scheres, S. H. W. (2017). Cryo-em structures of tau filaments from alzheimer’s disease. NATURE 547, 185+. doi:10.1038/nature23002.

4. Meisl, G., Xu, C. K., Taylor, J. D., Michaels, T. C. T., Levin, A., Otzen, D., Klenerman, D., Matthews, S., Linse, S., Andreasen, M., and Knowles, T. P. J. (2022). Uncovering the universality of self-replication in protein aggregation and its link to disease. Science Advances 8, eabn6831. URL: https://www.science.org/doi/abs/10.1126/sciadv.abn6831. doi:10.1126/sciadv.abn6831. arXiv:https://www.science.org/doi/pdf/10.1126/sciadv.abn6831.

5. Meisl, G. (2024). The thermodynamics of neurodegenerative disease. Biophysics Reviews 5, 011303. URL: https://doi.org/10.1063/5.0180899.doi:10.1063/5.0180899. arXiv:https://pubs.aip.org/aip/bpr/article-pdf/doi/10.1063/5.0180899/19837475/011303_1_5.0180899.pdf

6. Mudher, A., Colin, M., Dujardin, S., Medina, M., Dewachter, I., Naini, S. M. A., Mandelkow, E.-M., Mandelkow, E., Buee, L., Goedert, M., and Brion, J.-P. (2017). What is the evidence that tau pathology spreads through prion-like propagation? ACTA NEUROPATHOLOGICA COMMUNICATIONS 5. doi:10.1186/s40478-017-0488-7.

7. Rahayel, S., Zheng, Y.-Q., Liu, Z.-Q., Abdelgawad, A., Abbasi, N., Caputo, A., Zhang, B., Lo, A., Kehm, V., Kozak, M., Yoo, H. S., Dagher, A., and Luk, K. C. (2022). Differentially targeted seeding reveals unique pathological alpha-synuclein propagation patterns. BRAIN 145, 1743–1756. doi:10.1093/brain/awab440.

8. Vogel, J. W., Iturria-Medina, Y., Strandberg, O. T., Smith, R., Levitis, E., Evans, A. C., Hansson, O., Initiat, A. D. N., and Study, S. B. (2020). Spread of pathological tau proteins through communicating neurons in human alzheimer’s disease. NATURE COMMUNICATIONS 11. doi:10.1038/s41467-020-15701-2.

9. Clavaguera, F., Bolmont, T., Crowther, R. A., Abramowski, D., Frank, S., Probst, A., Fraser, G., Stalder, A. K., Beibel, M., Staufenbiel, M., Jucker, M., Goedert, M., and Tolnay, M. (2009). Transmission and spreading of tauopathy in transgenic mouse brain. NATURE CELL BIOLOGY 11, 909–U325. doi:10.1038/ncb1901.

10. Meisl, G., Hidari, E., Allinson, K., Rittman, T., DeVos, S. L., Sanchez, J. S., Xu, C. K., Duff, K. E., Johnson, K. A., Rowe, J. B., Hyman, B. T., Knowles, T. P. J., and Klenerman, D. (2021). In vivo rate-determining steps of tau seed accumulation in alzheimer’s disease. Science Advances 7, eabh1448. URL: https://www.science.org/doi/abs/10.1126/sciadv.abh1448. doi:10.1126/sciadv.abh1448. arXiv:https://www.science.org/doi/pdf/10.1126/sciadv.abh1448.

11. Dimou, E., Katsinelos, T., Meisl, G., Tuck, B. J., Keeling, S., Smith, A. E., Hidari, E., Lam, J. Y. L., Burke, M., Lovestam, S., Ranasinghe, R. T., McEwan, W. A., and Klenerman, D. (2023). Super-resolution imaging unveils the self-replication of tau aggregates upon seeding. CELL REPORTS 42. doi:10.1016/j.celrep.2023.112725.

12. Rollo, J., Crawford, J., and Hardy, J. (2023). A dynamical systems approach for multiscale synthesis of alzheimer’s pathogenesis. Neuron 111, 2126–2139. URL: https://www.sciencedirect.com/science/article/pii/S0896627323003033. doi:10.1016/j.neuron.2023.04.018.

13. Knowles, T. P. J., Waudby, C. A., Devlin, G. L., Cohen, S. I. A., Aguzzi, A., Vendruscolo, M., Terentjev, E. M., Welland, M. E., and Dobson, C. M. (2009). An analytical solution to the kinetics of breakable filament assembly. Science 326, 1533–1537. URL: https://www.science.org/doi/abs/10.1126/science.1178250. doi:10.1126/science.1178250. arXiv:https://www.science.org/doi/pdf/10.1126/science.1178250.

14. Meisl, G., Kirkegaard, J. B., Arosio, P., Michaels, T. C. T., Vendruscolo, M., Dobson, C. M., Linse, S., and Knowles, T. P. J. (2016). Molecular mechanisms of protein aggregation from global fitting of kinetic models. NATURE PROTOCOLS 11, 252–272. doi:10.1038/nprot.2016.010.

15. Meisl, G., Knowles, T. P. J., and Klenerman, D. (2022). Mechanistic models of protein aggregation across length-scales and time-scales: From the test tube to neurodegenerative disease. FRONTIERS IN NEUROSCIENCE 16. doi:10.3389/fnins.2022.909861.

16. Zheng, Y.-Q., Zhang, Y., Yau, Y., Zeighami, Y., Larcher, K., Misic, B., and Dagher, A. (2019). Local vulnerability and global connectivity jointly shape neurodegenerative disease propagation. PLOS BIOLOGY 17. doi:10.1371/journal.pbio.3000495.

17. Thompson, T. B., Chaggar, P., Kuhl, E., Goriely, A., and for the Alzheimer’s Disease Neuroimaging Initiative (2020). Protein-protein interactions in neurodegenerative diseases: A conspiracy theory. PLOS Computational Biology 16, 1–41. URL: https://doi.org/10.1371/journal.pcbi.1008267.doi:10.1371/journal.pcbi.1008267.

18. Raj, A., Kuceyeski, A., and Weiner, M. (2012). A network diffusion model of disease progression in dementia. NEURON 73, 1204–1215. doi:10.1016/j.neuron.2011.12.040.

19. Holland, N., Jones, P. S., Savulich, G., Wiggins, J. K., Hong, Y. T., Fryer, T. D., Manavaki, R., Sephton, S. M., Boros, I., Malpetti, M., Hezemans, F. H., Aigbirhio, F., I, Coles, J. P., O’Brien, J., and Rowe, J. B. (2020). Synaptic loss in primary tauopathies revealed by [¡sup¿11¡/sup¿c]ucb-jpositron emission tomography. MOVEMENT DISORDERS 35, 1834– 1842. doi:10.1002/mds.28188.

20. Cope, T. E., Rittman, T., Borchert, R. J., Jones, P. S., Vatansever, D., Allinson, K., Passamonti, L., Rodriguez, P. V., Bevan-Jones, W. R., O’Brien, J. T., and Rowe, J. B. (2018). Tau burden and the functional connectome in alzheimer’s disease and progressive supranuclear palsy. BRAIN 141, 550–567. doi:10.1093/brain/awx347.

21. Pantanowitz, L., Sharma, A., Carter, A. B., Kurc, T., Sussman, A., and Saltz, J. (2018). Twenty years of digital pathology: An overview of the road travelled, what is on the horizon, and the emergence of vendor-neutral archives. Journal of Pathology Informatics 9, 40. URL: https://www.sciencedirect.com/science/article/pii/S2153353922003534. 10.4103/jpi.jpi_69_18.

22. Pansuwan, T., Quaegebeur, A., Kaalund, S. S., Hidari, E., Briggs, M., Rowe, J. B., and Rittman, T. (2023). Accurate digital quantification of tau pathology in progressive supranuclear palsy. ACTA NEUROPATHOLOGICA COMMUNICATIONS 11. doi:10.1186/s40478-023-01674-y.

23. Martinez-Maldonado, A., Angel Ontiveros-Torres, M., Harrington, C. R., Francisco Montiel-Sosa, J., Garcia-Tapia Prandiz, R., Bocanegra-Lopez, P., Michael Sorsby-Vargas, A., Bravo-Munoz, M., Floran-Garduno, B., Villanueva-Fierro, I., Perry, G., Garces-Ramirez, L., de la Cruz, F., Martinez-Robles, S., Pacheco-Herrero, M., and Luna-Munoz, J. (2021). Molecular processing of tau protein in progressive supranuclear palsy: Neuronal and glial degeneration. JOURNAL OF ALZHEIMERS DISEASE 79, 1517–1531. doi:10.3233/JAD-201139.

24. Kovacs, G. G., Lukic, M. J., Irwin, D. J., Arzberger, T., Respondek, G., Lee, E. B., Coughlin, D., Giese, A., Grossman, M., Kurz, C., McMillan, C. T., Gelpi, E., Compta, Y., van Swieten, J. C., Donker-Kaat, L., Troakes, C., Al-Sarraj, S., Robinson, J. L., Roeber, S., Xie, S. X., Lee, V. M.-Y., Trojanowski, J. Q., and Hoeglinger, G. U. (2020). Distribution patterns of tau pathology in progressive supranuclear palsy. ACTA NEUROPATHOLOGICA 140, 99–119. doi:10.1007/s00401-020-02158-2.

25. Rodriguez Camargo, D. C., Sileikis, E., Chia, S., Axell, E., Bernfur, K., Cataldi, R. L., Cohen, S. I. A., Meisl, G., Habchi, J., Knowles, T. P. J., Vendruscolo, M., and Linse, S. (2021). Proliferation of tau 304–380 fragment aggregates through autocatalytic secondary nucleation. ACS Chemical Neuroscience 12, 4406–4415. URL: https://doi.org/10.1021/acschemneuro.1c00454.doi:10.1021/acschemneuro.1c00454. arXiv:https://doi.org/10.1021/acschemneuro.1c00454.PMID: 34783519.

26. Thompson, T. B., Meisl, G., Knowles, T. P. J., and Goriely, A. (2021). The role of clearance mechanisms in the kinetics of pathological protein aggregation involved in neurodegenerative diseases. The Journal of Chemical Physics 154, 125101. URL: https://doi.org/10.1063/5.0031650.doi:10.1063/5.0031650. arXiv:https://pubs.aip.org/aip/jcp/article-pdf/doi/10.1063/5.0031650/14015306/1

27. Andrews, R., Fu, B., Toomey, C. E., Breiter, J. C., Lachica, J., Tian, R., Beckwith, J. S., Needham, L.-M., Chant, G. J., Loiseau, C., Deconfin, A., Baspin, K., Magill, P. J., Jaunmuktane, Z., Freeman, O. J., Taylor, B. J. M., Hardy, J., Lashley, T., Ryten, M., Vendruscolo, M., Wood, N. W., Weiss, L. E., Gandhi, S., and Lee, S. F. (2024). Large-scale visualisation of α-synuclein oligomers in parkinson’s disease brain tissue. bioRxiv. URL: https://www.biorxiv.org/content/early/2024/02/19/2024.02.17.580698. doi:10.1101/2024.02.17.580698. arXiv:https://www.biorxiv.org/content/early/2024/02/19/2024.02.17.580698.full.pdf

28. Akiyama, H., Barger, S., Barnum, S., Bradt, B., Bauer, J., Cole, G. M., Cooper, N. R., Eikelenboom, P., Emmerling, M., Fiebich, B. L., Finch, C. E., Frautschy, S., Griffin, W. S., Hampel, H., Hull, M., Landreth, G., Lue, L., Mrak, R., Mackenzie, I. R., McGeer, P. L., O’Banion, M. K., Pachter, J., Pasinetti, G., Plata-Salaman, C., Rogers, J., Rydel, R., Shen, Y., Streit, W., Strohmeyer, R., Tooyoma, I., Van Muiswinkel, F. L., Veerhuis, R., Walker, D., Webster, S., Wegrzyniak, B., Wenk, G., and Wyss-Coray, T. (2000). Inflammation and alzheimer’s disease. Neurobiology of aging 21, 383–421. URL: https://www.ncbi.nlm.nih.gov/pubmed/10858586. doi:10.1016/s0197-4580(00)00124-x.

29. Fu, H., Hardy, J., and Duff, K. E. (2018). Selective vulnerability in neurodegenerative diseases. NATURE NEUROSCIENCE 21, 1350–1358. doi:10.1038/s41593-018-0221-2.

30. Praschberger, R., Kuenen, S., Schoovaerts, N., Kaempf, N., Singh, J., Janssens, J., Swerts, J., Nachman, E., Calatayud, C., Aerts, S., Poovathingal, S., and Verstreken, P. (2023). Neuronal identity defines α-synuclein and tau toxicity. Neuron 111, 1577–1590.e11. URL: https://www.sciencedirect.com/science/article/pii/S0896627323001666. 10.1016/j.neuron.2023.02.033.

31. Fu, H., Possenti, A., Freer, R., Nakano, Y., Villegas, N. C. H., Tang, M., Cauhy, P. V. M., Lassus, B. A., Chen, S., Fowler, S. L., Figueroa, H. Y., Huey, E. D., Johnson, G. V. W., Vendruscolo, M., and Duff, K. E. (2019). A tau homeostasis signature is linked with the cellular and regional vulnerability of excitatory neurons to tau pathology. NATURE NEUROSCIENCE 22, 47+. doi:10.1038/s41593-018-0298-7.

32. Tardivel, M., Bégard, S., Bousset, L., Dujardin, S., Coens, A., Melki, R., Buée, L., and Colin, M. (2016). Tunneling nanotube (TNT)-mediated neuron-to neuron transfer of pathological Tau protein assemblies. Acta Neuropathologica Communications 4, 117. doi:10.1186/s40478-016-0386-4.

33. Dickson, D. W., Rademakers, R., and Hutton, M. L. (2007). Progressive supranuclear palsy: Pathology and genetics. BRAIN PATHOLOGY 17, 74–82. doi:10.1111/j.1750-3639.2007.00054.x.

34. Morimoto, R. I. (2020). Cell-nonautonomous regulation of proteostasis in aging and disease. Cold Spring Harbor Perspectives in Biology 12. URL: http://cshperspectives.cshlp.org/content/12/4/a034074.abstract. doi:10.1101/cshperspect.a034074. arXiv:http://cshperspectives.cshlp.org/content/12/4/a034074.full.pdf+html.

35. Hindle, J. V. (2010). Ageing, neurodegeneration and parkinson’s disease. Age and Ageing 39, 156–161. URL: https://doi.org/10.1093/ageing/afp223.doi:10.1093/ageing/afp223. arXiv:https://academic.oup.com/ageing/article-pdf/39/2/156/88338/afp223.pdf.

36. Labbadia, J., and Morimoto, R. I. (2015). The biology of proteostasis in aging and disease. Annual Review of Biochemistry 84, 435–464. URL: https://doi.org/10.1146/annurev-biochem-060614-033955.doi:10.1146/annurev-biochem-060614-033955. arXiv:https://doi.org/10.1146/annurev-biochem-060614-033955.

37. Benveniste, H., Liu, X., Koundal, S., Sanggaard, S., Lee, H., and Wardlaw, J. (2019). The glymphatic system and waste clearance with brain aging: A review. Gerontology 65, 106–119. URL: https://doi.org/10.1159/000490349.doi:10.1159/000490349. arXiv:https://karger.com/ger/article-pdf/65/2/106/2839264/000490349.pdf.

38. Saramä ki, J., and Kaski, K. (2005). Modelling development of epidemics with dynamic small-world networks. JOURNAL OF THEORETICAL BIOLOGY 234, 413–421. doi:10.1016/j.jtbi.2004.12.003.

39. Shafiei, G., Bazinet, V., Dadar, M., Manera, A. L., Collins, D. L., Dagher, A., Borroni, B., Sanchez-Valle, R., Moreno, F., Laforce, R., Graff, C., Synofzik, M., Galimberti, D., Rowe, J. B., Masellis, M., Tartaglia, M. C., Finger, E., Vandenberghe, R., de Mendonca, A., Tagliavini, F., Santana, I., Butler, C., Gerhard, A., Danek, A., Levin, J., Otto, M., Sorbi, S., Jiskoot, L. C., Seelaar, H., van Swieten, J. C., Rohrer, J. D., Misic, B., Ducharme, S., Degeneration, F. L., and In, G. F. D. (2023). Network structure and transcriptomic vulnerability shape atrophy in frontotemporal dementia. BRAIN 146, 321–336. doi:10.1093/brain/awac069.

40. Kröger, M., and Schlickeiser, R. (2020). Analytical solution of the sir-model for the temporal evolution of epidemics. part a: time-independent reproduction factor. Journal of Physics A: Mathematical and Theoretical 53, 505601. URL: https://dx.doi.org/10.1088/1751-8121/abc65d. doi:10.1088/1751-8121/abc65d.

41. Knowles, T. P. J., White, D. A., Abate, A. R., Agresti, J. J., Cohen, S. I. A., Sperling, R. A., Genst, E. J. D., Dobson, C. M., and Weitz, D. A. (2011). Observation of spatial propagation of amyloid assembly from single nuclei. Proceedings of the National Academy of Sciences 108, 14746–14751. URL: https://www.pnas.org/doi/abs/10.1073/pnas.1105555108. doi:10.1073/pnas.1105555108. arXiv:https://www.pnas.org/doi/pdf/10.1073/pnas.1105555108.

42. Weickenmeier, J., Kuhl, E., and Goriely, A. (2018). Multiphysics of prionlike diseases: Progression and atrophy. Phys. Rev. Lett. 121, 158101. URL: https://link.aps.org/doi/10.1103/PhysRevLett.121.158101. doi:10.1103/PhysRevLett.121.158101.

43. Frost, B., Jacks, R. L., and Diamond, M. I. (2009). Propagation of tau misfolding from the outside to the inside of a cell*. Journal of Biological Chemistry 284, 12845–12852. URL: https://www.sciencedirect.com/science/article/pii/S0021925820582847. 10.1074/jbc.M808759200.

44. Grothe, M. J., Sepulcre, J., Gonzalez-Escamilla, G., Jelistratova, I., Schöll, M., Hansson, O., Teipel, S. J., and Initiative, A. D. N. (2018). Molecular properties underlying regional vulnerability to Alzheimer’s disease pathology. Brain 141, 2755–2771. URL: https://doi.org/10.1093/brain/awy189.doi:10.1093/brain/awy189. arXiv:https://academic.oup.com/brain/article-pdf/141/9/2755/25590486/awy189.pdf.

45. Andrade-Moraes, C. H., Oliveira-Pinto, A. V., Castro-Fonseca, E., da Silva, C. G., Guimaräes, D. M., Szczupak, D., Parente-Bruno, D. R., Carvalho, L. R., Polichiso, L., Gomes, B. V., Oliveira, L. M., Rodriguez, R. D., Leite, R. E., Ferretti-Rebustini, R. E., Jacob-Filho, W., Pasqualucci, C. A., Grinberg, L. T., and Lent, R. (2013). Cell number changes in Alzheimer’s disease relate to dementia, not to plaques and tangles. Brain 136, 3738–3752. URL: https://doi.org/10.1093/brain/awt273.doi:10.1093/brain/awt273. arXiv:https://academic.oup.com/brain/article-pdf/136/12/3738/13795085/awt273.pdf

46. Farrell, K., Humphrey, J., Chang, T., Zhao, Y., Leung, Y. Y., Kuksa, P. P., Patil, V., Lee, W.-P., Kuzma, A. B., Valladares, O., Cantwell, L. B., Wang, H., Ravi, A., De Sanctis, C., Han, N., Christie, T. D., Afzal, R., Kandoi, S., Whitney, K., Krassner, M. M., Ressler, H., Kim, S., Dangoor, D., Iida, M. A., Casella, A., Walker, R. H., Nirenberg, M. J., Renton, A. E., Babrowicz, B., Coppola, G., Raj, T., Höglinger, G. U., Müller, U., Golbe, L. I., Morris, H. R., Hardy, J., Revesz, T., Warner, T. T., Jaunmuktane, Z., Mok, K. Y., Rademakers, R., Dickson, D. W., Ross, O. A., Wang, L.-S., Goate, A., Schellenberg, G., Geschwind, D. H., Hopfner, F., Roeber, S., Herms, J., Troakes, C., Gelpi, E., Compta, Y., van Swieten, J. C., Rajput, A., Hinton, F., de Yebenes, J. G., Crary, J. F., Naj, A., and Group, P. G. S. (2024). Genetic, transcriptomic, histological, and biochemical analysis of progressive supranuclear palsy implicates glial activation and novel risk genes. Nature Communications 15, 7880. URL: https://doi.org/10.1038/s41467-024-52025-x.doi:10.1038/s41467-024-52025-x.

